# SynthCraft: an AI partner for synthetic data generation to support data access and augmentation in healthcare

**DOI:** 10.1101/2025.08.17.25333866

**Authors:** Thomas Callender, Anders Boyd, Robert Davis, Silas Ruhrberg Estevez, Juan Lavista Ferres, Mihaela van der Schaar

**Affiliations:** Department of Public Health and Primary Care, University of Cambridge, United Kingdom; Cambridge Centre for AI in Medicine, University of Cambridge, United Kingdom; Department of Infectious Diseases, Inselspital, Bern University Hospital, University of Bern, Switzerland; Amsterdam UMC location University of Amsterdam, Department of Infectious Diseases, Meibergdreef 9, Amsterdam, the Netherlands; Amsterdam Institute for Immunology and Infectious Diseases, Infectious Diseases, Amsterdam, the Netherlands; School of Clinical Medicine, University of Cambridge, United Kingdom; Department of Applied Mathematics and Theoretical Physics, University of Cambridge, United Kingdom; AI for Good Lab, Microsoft, Redmond, Washington, United States of America

## Abstract

**Background:** Access to high-quality data provides the foundation for biomedical research. But data access is often limited or challenging due to privacy constraints, whilst the data themselves may be unrepresentative or sparse. Synthetic data can support privacy-preserving data access, data augmentation, as well as complex analytical workflows for the development of digital twins or to evaluate the impacts of data distribution shifts. However, the use of synthetic data remains limited due to the complexity of the methods themselves and their evaluation, as well as the need for advanced programming skills.

**Methods:** We developed SynthCraft, a tool for AI-human collaboration to support the principled, transparent, use of state-of-the-art synthetic data generation methods. SynthCraft uses Large Language Models (LLMs) combined with a reinforcement learning-based reasoning engine to orchestrate the necessary workflow to generate synthetic data based on dynamic interaction with the user using natural language. We demonstrate the capability of SynthCraft with both tabular and genomic datasets: National Health and Nutrition Examination Survey (NHANES) and the Cancer Genome Atlas (TCGA).

**Results:** Using SynthCraft, we analysed the privacy, statistical fidelity, and downstream utility of four different synthetic data generators both with and without explicit privacy-preserving designs when applied to both the NHANES and TCGA datasets. We show that how different generators perform differently – and that no single method was optimal – across varying use-cases and datasets. Furthermore, we demonstrate how SynthCraft can be used for data augmentation as part of a workflow to attempt to mitigate imbalances in the proportion of individuals from different ethnic backgrounds.

**Conclusions:** An LLM-based, human-in-the-loop, AI partner can support the generation of synthetic datasets. Such tools could improve the quality, reproducibility, and transparency of research methods, whilst increasing their accessibility. Research into their use across different methodological areas is warranted.

## INTRODUCTION

Healthcare research is premised on access to high-quality data. Nevertheless, legitimate privacy concerns mean healthcare data are often difficult to access if not entirely unavailable^1^. When available, data may be sparse, subject to biases, or unrepresentative^2^. The implications of this are felt throughout biomedical research.

Synthetic data has emerged as a powerful approach to overcome these problems. Rather than masking or anonymizing real data, synthetic data are generated to mirror the statistical patterns and relationships found in real datasets^2,3^. Because of this, synthetic data can support privacy-preserving data access and the development of digital twins^2–5^. When applied with care, synthetic data can also play a role in mitigating issues of fairness, biases, and data sparsity through data augmentation and adaptation^2,4^. While the benefits of synthetic data are increasingly recognised, its application in healthcare remains in its infancy^6^. Key challenges include the complexity of developing synthetic data, the speed with which synthetic data methodologies are being developed, and the lack of standardisation of the metrics by which synthetic data should be judged^7,8^.

Efforts to improve the quality and accessibility of research methods have historically revolved around training, the development of software packages that abstract elements of how a method is implemented^9–11^, and reporting guidelines^12^. Software for synthetic data have begun to bridge this gap^9–11^, yet their use still demands advanced programming and familiarity with complex data pipelines; skills that many healthcare researchers do not possess or lack the time to apply effectively^13^. Guidelines have been used as both an educational tool and to improve the quality of research^12^, but their impact has been mixed^14^.

Here we introduce SynthCraft, a large language model (LLM)-based partner for synthetic data development as a solution to these barriers. The system use LLMs to orchestrate multiple sequential or concurrent steps to solve complex problems. Working together entirely in natural language, SynthCraft empowers a researcher with state-of-the-art synthetic data software libraries to build synthetic data by progressing through a principled, stepwise, approach, discussing problems and solutions as they arise. We demonstrate the capability of SynthCraft across tabular and genomic datasets for both data access and augmentation tasks.

## METHODS

### Overview of SynthCraft

Designed to act as a “human-in-the-loop” partner, SynthCraft guides users through an interactive, step-by-step process (Figure 1). First, SynthCraft characterises the real dataset, identifying key features and the data structure, as well as performing exploratory data analyses. SynthCraft then engages the user to consider the most appropriate synthetic data generation methods for their circumstances, explaining the different strengths and weakness of alternative approaches, before invoking Synthcity to generate synthetic data itself^9^.

**Figure 1:**
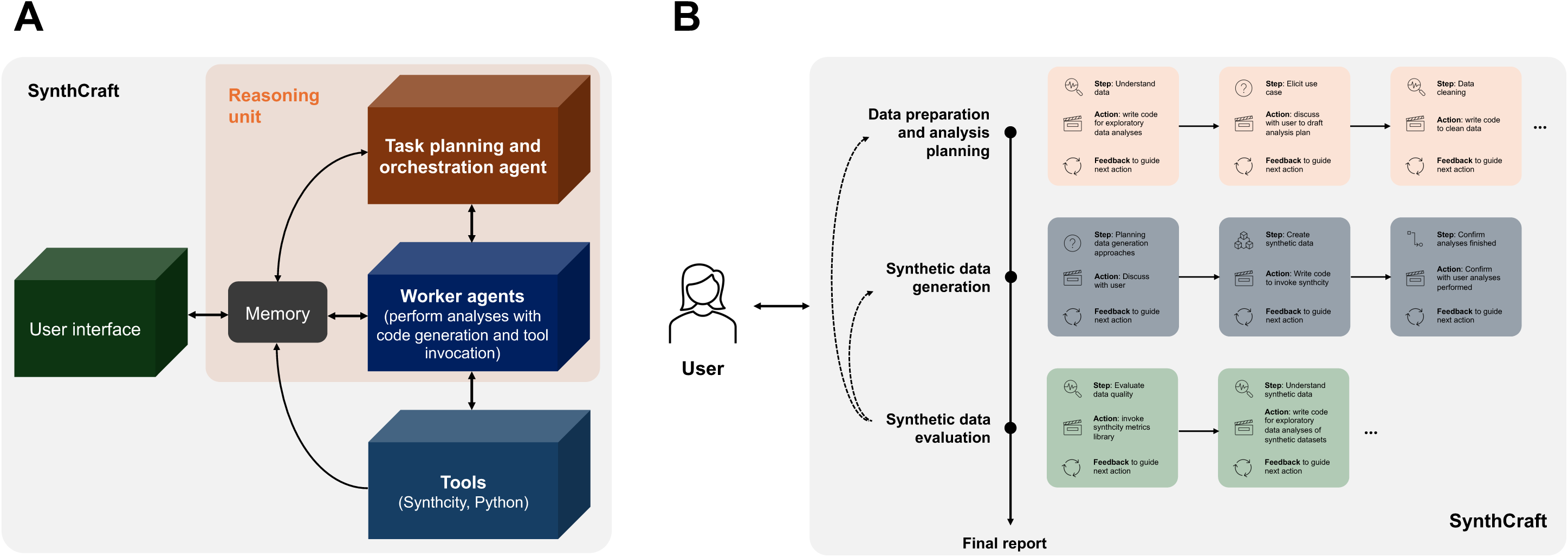
Overview of SynthCraft. **(A)** SynthCraft is a modular framework consisting of large language model (LLM) agents with access to tools (SynthCity, Python) linked with working memory and underpinned by a reasoning system for in-context learning. Interaction with the user is through a natural language interface. An illustration of SynthCraft’s synthetic data generation pipeline is shown in (**B**) alongside a schematic of how the episodic multi-armed bandit reasoning approach works (to the right of the vertical line)^15,16^. At each stage of the pipeline, from data preparation and developing an analysis plan through evaluating the quality of the synthetic data generated, multiple intermediate steps may be needed. In the illustration, we have simplified for clarity, but at each step the agent has a particular state, reflecting progress towards completing the relevant task, can select appropriate actions, and then receives feedback either from interaction with the user, external tools invoked, or self-reflection^15,16^. The number of episodes – or steps – will vary depending on the use case, the dataset, and the interaction with the user. Though presented as a sequential pipeline, SynthCraft may need to return to earlier stages; for example, to trial alternative synthetic data generators if the quality of the synthetic data generated for a given task is insufficient at the evaluation stage. Subfigures adapted from refs ^15,16^.

Synthcity is an open-source package with extensive community engagement that provides a standardised interface for accessing and evaluating a comprehensive array of generators for any synthetic data use case – from synthetic data generation to managing privacy, fairness, domain adaptation and image generation^9^ (Supplementary Table S1). Together, the user and partner compare the generated data and iteratively refine analyses based on user feedback. The process has been designed to minimise ambiguity, ensure methodological rigour, and quality-assure the generated synthetic data. On completion, a structured report detailing all steps taken, decisions made, and code run, is produced to support transparency and reproducibility.

SynthCraft is a modular LLM framework, in which a coordinating LLM agent controls worker agents with the ability to generate code and run specialised tools – in this case, Synthcity. We used GPT-4 from OpenAI (model: gpt-40-2024-05-13; API version: “2024-02-01”; temperature: 0.5) on a secured instance of Azure as the underlying LLM in these analyses, but an alternative LLM could be simply interchanged to underpin the agents in this framework. The LLMs themselves are not privy to the real or synthetic data at any point, only prompts – the instructions provided by the user – whilst all analyses are undertaken on a user’s own device.

The collaborative, sequential, decision-making process between the user and SynthCraft uses a reasoning system trained using multi-armed bandits, a type of reinforcement learning designed for complex multi-stage optimisation problems (Figure 1)^15,16^. Each stage in the process by which synthetic data are generated – which are encoded as fixed logic – can be considered as an episode consisting of one or more steps. The steps necessary for each episode will develop based on the needs of the user and characteristics of the data – i.e., adaptive learning^15^. For example, because the performance of synthetic data generation methods differs between use-cases and datasets, the generative step may require several iterative cycles before completion. At each step, an action is taken, such as running a synthetic data generator tool or producing a report. Each action is associated with both a cost and a reward. Should the task be completed or a problem encountered by the LLM that requires discussion with the user, a stop action occurs. Feedback received from all other actions taken within that episode informs the next action to be taken. The overall objective of the LLM is to maximise net rewards (total rewards less costs), which occurs on successful completion of the task. Further details are presented in the Supplementary Methods.

### Datasets

To demonstrate the capacity of SynthCraft to generate synthetic datasets across both epidemiological data and genomic data, we used National Health and Nutrition Examination Survey (NHANES) (wave 2021-2023)^17^ and The Cancer Genome Atlas (TCGA)^18^. In brief, the NHANES study is a complex, stratified, multistage cluster probability sample of the civilian, noninstitutionalized population of the United States of America (USA). NHANES collects data through household interviews (for demographics, diet, tobacco use, and medical history) and a mobile examination centre (for health, dental, anthropometric, and biochemical examinations and biospecimen collection). For this study, we included the covariates age, gender, ethnicity, body mass index (BMI), waist circumference, total cholesterol levels, insulin levels, and self-reported myocardial infarction. We excluded participants who had missing data on these covariates, resulting in a dataset of 2,924 unique observations.

The Cancer Genome Atlas (TCGA) reflects the high-dimensional, heterogeneous data structures characteristic of multi-omics studies. TCGA is an ongoing collaboration between the National Cancer Institute and National Human Genome Research Institute in the USA with the aim of generating comprehensive, multi-dimensional maps of the key genomic changes in 33 types of cancer^18^. TCGA contains multi-omic data on tumour tissue and matched normal tissues from more than 11,000 patients alongside clinical information. For this study, we included data on tumour purity in bulk RNA sequencing (using the Illumina HiSeq 2000 platform), as determined from the ABSOLUTE algorithm^19^ and a previously derived gene signature^20^. We included all tumour samples for which there were no missing values in gene expression, resulting in a dataset of 9,678 samples.

### Synthetic data generation and performance evaluation

We generated synthetic versions of both the NHANES and TCGA datasets using the following four data generators, selected to represent a range of generators both with and without specific privacy-preserving features, trained with their default hyperparameters: Private Aggregation of Teacher Ensembles Generative Adversarial Networks (PATE-GAN)^21^, Denoising diffusion probabilistic models (DDPM)^22^, Anonymization through Data Synthesis using Generative Adversarial Networks (ADSGAN)^23^, and Conditional Tabular Generative Adversarial Network (CT-GAN)^24^. PATE-GAN, DDPM, and ADSGAN are specifically privacy-preserving synthetic data generators. PATE-GAN uses differential privacy, a mathematical framework that ensures the inclusion or exclusion of a single data point does not significantly affect the output of data analysis^25^. We used a default privacy epsilon of 1 for these analyses. DDPM and ADS-GAN are specifically tailored to protect against re-identification attacks, where an adversary attempts to identify individuals within anonymised or pseudonymised datasets. CT-GAN has no additional privacy-preserving framework embedded.

To evaluate the quality, utility, and privacy of the generated data, we focused on several performance metrics (an exhaustive list of metrics and results are presented in Supplementary Table S2)^9,26,27^. For data quality, we used several metrics including Jensen-Shannon distance, the empirical maximum mean discrepancy, and α-precision^26^. To evaluate the utility of the data for downstream tasks, we then built predictive models and measured the MSE (for regression) and the area under the received operating characteristic (AUC) curve (for classification) on both training and out-of-distribution data. For data privacy, we measured its authenticity, k-anonymity, and identifiability score. Authenticity quantifies the percentage of generated samples that are not near-identical copies of any real training example^26^. K-anonymization involves quantifying the smallest k such that, in the real data (for ground truth) or the synthetic data, every combination of quasi-identifiers appears at least k times. The identifiability score assesses how easily a synthetic record can be linked back to a specific real individual by comparing quasi-identifiers between real and synthetic datasets to test for privacy in the synthetic dataset.

### Synthetic data augmentation

In common with many research cohorts, NHANES does not have equal representation across different ethnicities. To address this, we used a four-stage augmentation pipeline: baseline enumeration; enrichment target calculation; conditional synthetic generation; and cohort integration and evaluation. This allowed us to train models on augmented datasets containing the same number of cases from each ethnic group represented. Further details can be found in the Supplementary Methods.

### Statistical analysis

To compare distributions of variables between real and synthetic datasets, we calculated the counts and percentages (for categorical variables) and medians and interquartile ranges (IQR) (for continuous variables) in the real and synthetic datasets in the NHANES dataset and mean and standard deviation for the gene expression in the TCGA dataset.

For the real and synthetic NHANES databases, we modelled the relationship between self-reported myocardial infarction using logistic regression and including all other variables in the dataset as covariables. We obtained odds ratios (OR) and 95% confidence intervals (CI) from these models. We subsequently built prediction models, analysing discriminative performance (area under the receiver operating curve; AUC) in aggregate and across sub-groups using bootstrapped (1,000 runs) confidence intervals. For the TCGA dataset, we calculated the mean square error (MSE) of the predicted purity. We used a 5-fold cross-validation with 80% training and 20% testing data in each fold reporting the mean error across the folds alongside the standard deviation.

## RESULTS

SynthCraft used chain-of-thought reasoning to adapt its workflow to generate high-quality synthetic versions of both a tabular epidemiological dataset - NHANES - and a genomic dataset - TCGA (Figure 1). A complete example of this workflow can be found in Supplementary Figure S1.

First, we compared the quality of the different datasets generated to the original NHANES dataset. The synthetic datasets generated with CT-GAN, DDPM, and ADS-GAN had similar statistical measures of fidelity to each other (Supplementary Table S3), whilst replicating the variable distributions seen in the original dataset (Table 1). Despite consistency at an aggregate level, all three models showed some departure from the original dataset in the proportion of individuals with the outcome of interest -myocardial infarction - with 2.5%, 3.0%, and 7.4% of the synthetic cohorts generated using CT-GAN, ADS-GAN, and DDPM, respectively, having the outcome relative to 4.2% of the real cohort. By contrast, the data generated by PATE-GAN had both lower statistical measures of fidelity and more divergence from the descriptive characteristics of the NHANES data, but would be considered more private, with greater k-anonymity and authenticity metrics, and the lowest identifiability scores (Supplementary Table S3).

**Table 1.**
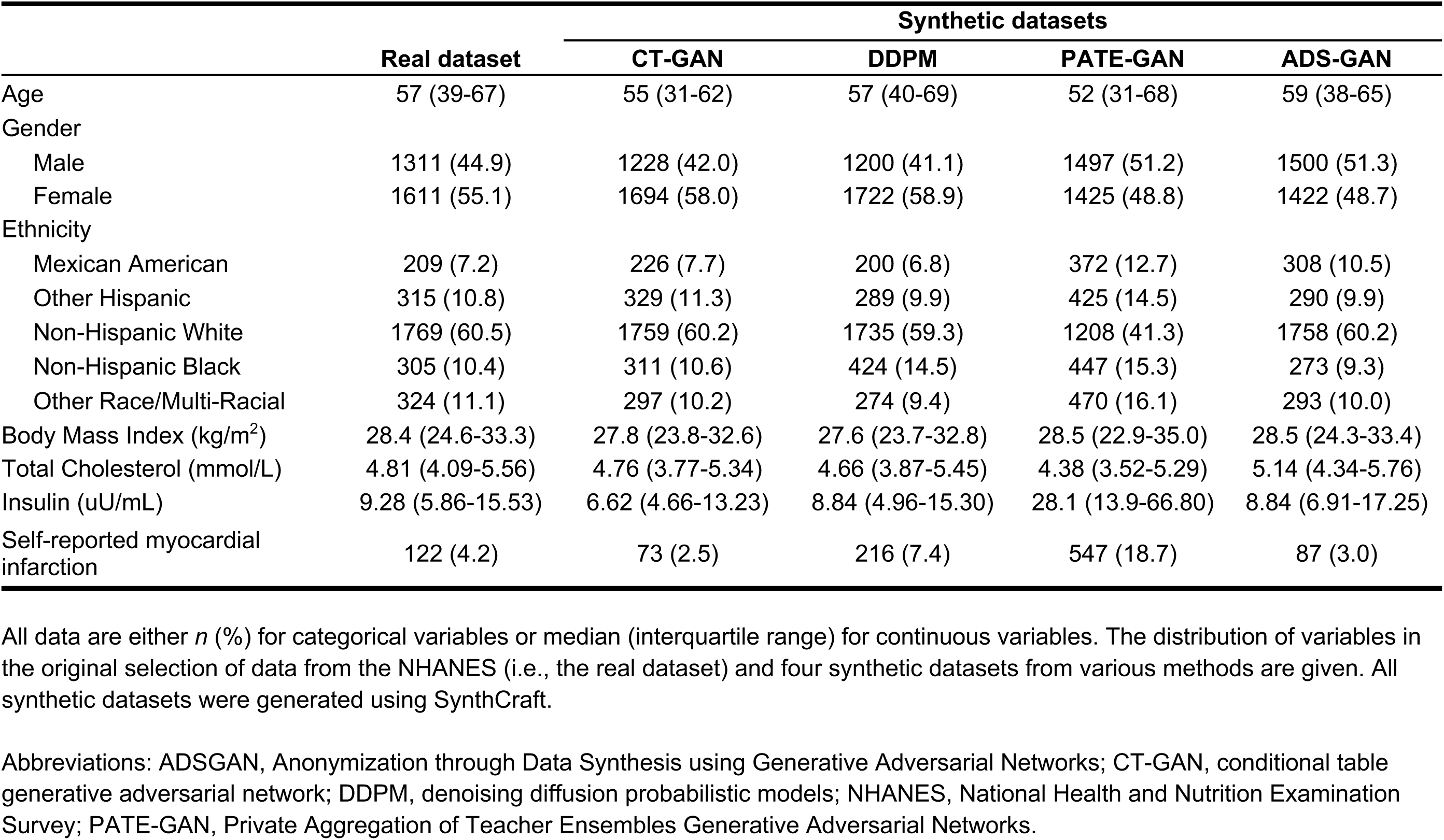
Comparison of variable distributions in the real and synthetically generated datasets (NHANES dataset)

|  | Real dataset | Synthetic datasets |  |  |  |
| --- | --- | --- | --- | --- | --- |
|  |  | CT-GAN | DDPM | PATE-GAN | ADS-GAN |
| Age | 57 (39-67) | 55 (31-62) | 57 (40-69) | 52 (31-68) | 59 (38-65) |
| Gender |  |  |  |  |  |
| Male | 1311 (44.9) | 1228 (42.0) | 1200 (41.1) | 1497 (51.2) | 1500 (51.3) |
| Female | 1611 (55.1) | 1694 (58.0) | 1722 (58.9) | 1425 (48.8) | 1422 (48.7) |
| Ethnicity |  |  |  |  |  |
| Mexican American | 209 (7.2) | 226 (7.7) | 200 (6.8) | 372 (12.7) | 308 (10.5) |
| Other Hispanic | 315 (10.8) | 329 (11.3) | 289 (9.9) | 425 (14.5) | 290 (9.9) |
| Non-Hispanic White | 1769 (60.5) | 1759 (60.2) | 1735 (59.3) | 1208 (41.3) | 1758 (60.2) |
| Non-Hispanic Black | 305 (10.4) | 311 (10.6) | 424 (14.5) | 447 (15.3) | 273 (9.3) |
| Other Race/Multi-Racial | 324 (11.1) | 297 (10.2) | 274 (9.4) | 470 (16.1) | 293 (10.0) |
| Body Mass Index (kg/m<sup>2</sup>) | 28.4 (24.6-33.3) | 27.8 (23.8-32.6) | 27.6 (23.7-32.8) | 28.5 (22.9-35.0) | 28.5 (24.3-33.4) |
| Total Cholesterol (mmol/L) | 4.81 (4.09-5.56) | 4.76 (3.77-5.34) | 4.66 (3.87-5.45) | 4.38 (3.52-5.29) | 5.14 (4.34-5.76) |
| Insulin (uU/mL) | 9.28 (5.86-15.53) | 6.62 (4.66-13.23) | 8.84 (4.96-15.30) | 28.1 (13.9-66.80) | 8.84 (6.91-17.25) |
| Self-reported myocardial infarction | 122 (4.2) | 73 (2.5) | 216 (7.4) | 547 (18.7) | 87 (3.0) |

All data are either $n$ (%) for categorical variables or median (interquartile range) for continuous variables. The distribution of variables in the original selection of data from the NHANES (i.e., the real dataset) and four synthetic datasets from various methods are given. All synthetic datasets were generated using SynthCraft.
Abbreviations: ADSSGAN, Anonymization through Data Synthesis using Generative Adversarial Networks; CT-GAN, conditional table generative adversarial network; DDPM, denoising diffusion probabilistic models; NHANES, National Health and Nutrition Examination Survey; PATE-GAN, Private Aggregation of Teacher Ensembles Generative Adversarial Networks.

We then modelled the relationship between the covariates and the occurrence of myocardial infarction in each dataset using logistic regression (Table 2). In keeping with the statistical measures of fidelity and descriptive statistics, regression models generated using synthetic data broadly preserved the relationships between variables and outcome seen in the real dataset. CT-GAN reproduced the parameter estimates found in the real dataset most closely, however there was a notable inversion in the relationship between female sex and myocardial infarction (13% increased risk amongst CT-GAN-generated synthetic data compared to a 32% reduction in risk in the real dataset).

**Table 2.**
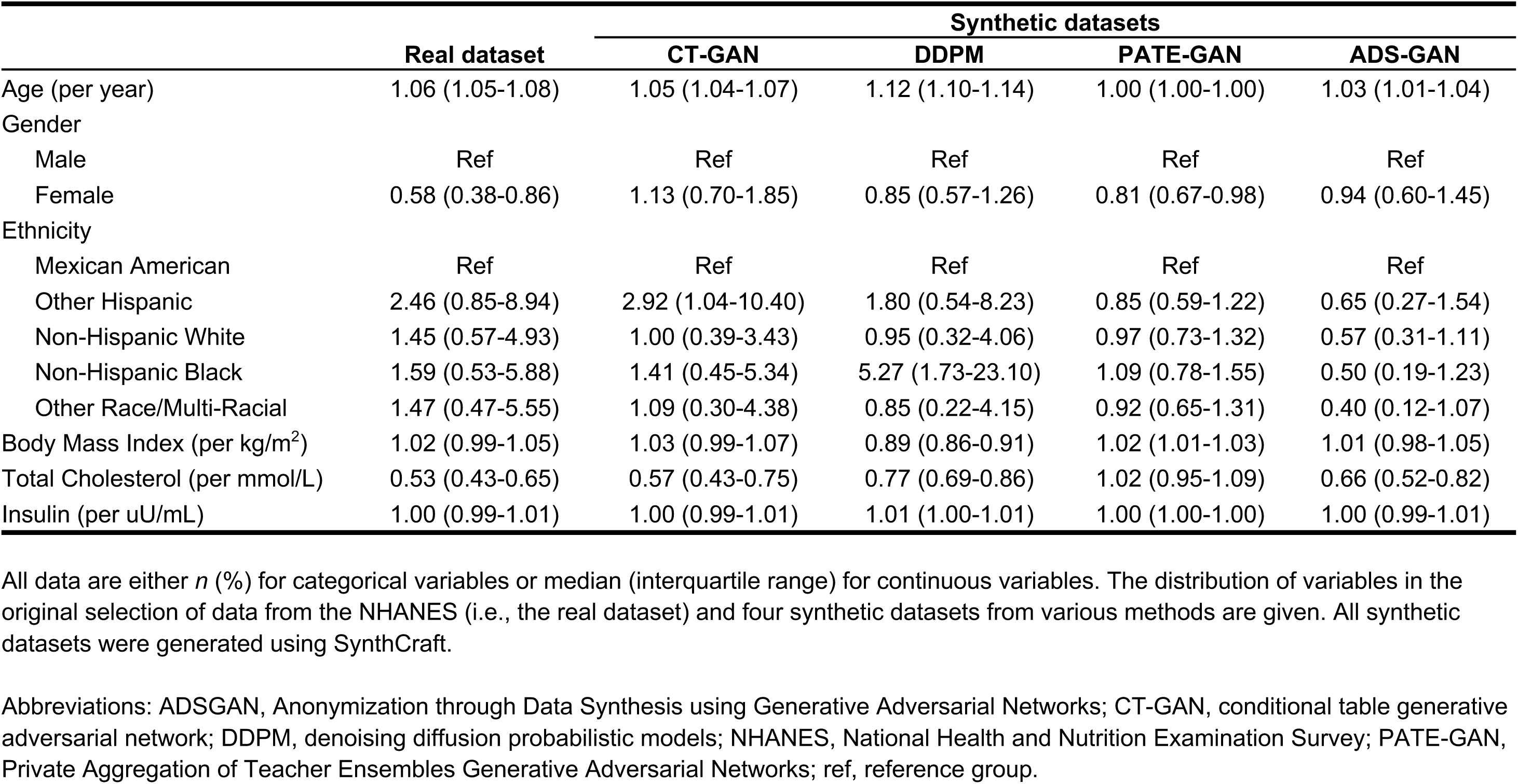
Comparison of regression parameters for self-reported myocardial infarction estimates in the real and synthetically generated datasets (NHANES dataset)

We subsequently analysed the discriminative performance of logistic regression models trained on synthetic data when tested on real data (Figure 2). Models trained on purely synthetic data generated using ADSGAN and CTGAN showed near equivalent AUCs to a model trained on the original dataset (real: 0.826, 95% confidence intervals [CI]: 0.785-0.864; ADSGAN: 0.810, 95% CI: 0.765-0.852; CTGAN: 0.802 95% CI: 0.757-0.845), although models trained on PATEGAN and DDPM performed less well. These patterns were maintained when analysing performance by age and ethnicity sub-groups (Figure 3).

**Figure 2:**
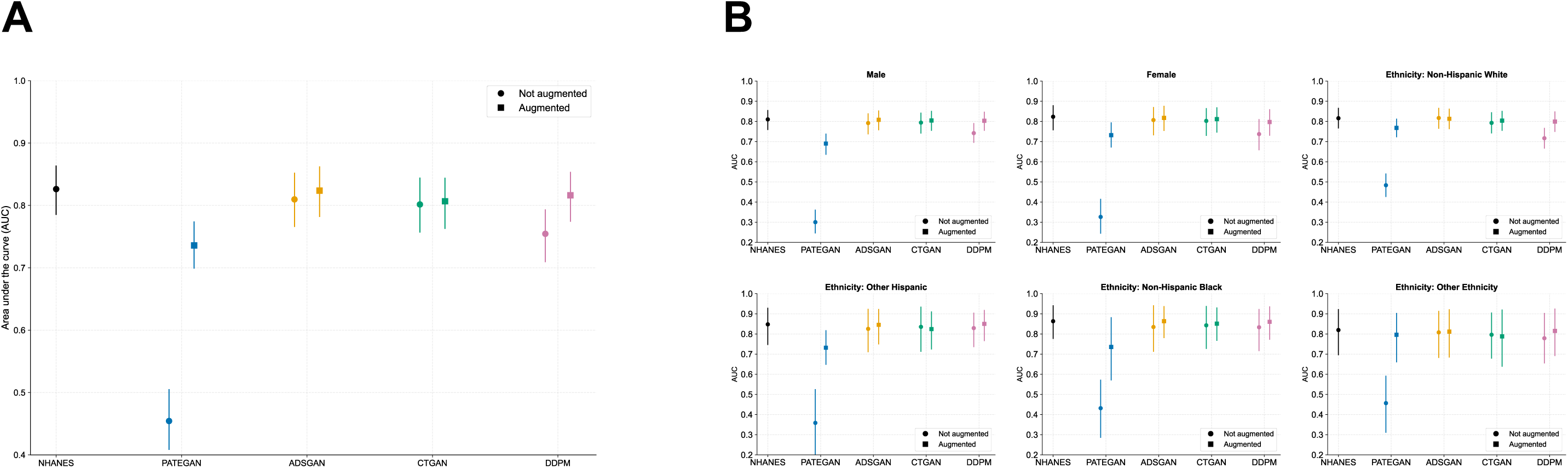
Discriminative performance (AUC) overall (A) and by sub-group (B) of logistic regression models trained on the original (NHANES) dataset, purely synthetic datasets (circles), or original dataset with augmentation (squares) when tested on the original real dataset. Abbreviations: ADSGAN, Anonymization through Data Synthesis using Generative Adversarial Networks; CT-GAN, conditional table generative adversarial network; DDPM, denoising diffusion probabilistic models; NHANES, National Health and Nutrition Examination Survey; PATEGAN, Private Aggregation of Teacher Ensembles Generative Adversarial Networks; AUC, area under the curve.

**Figure 3:**
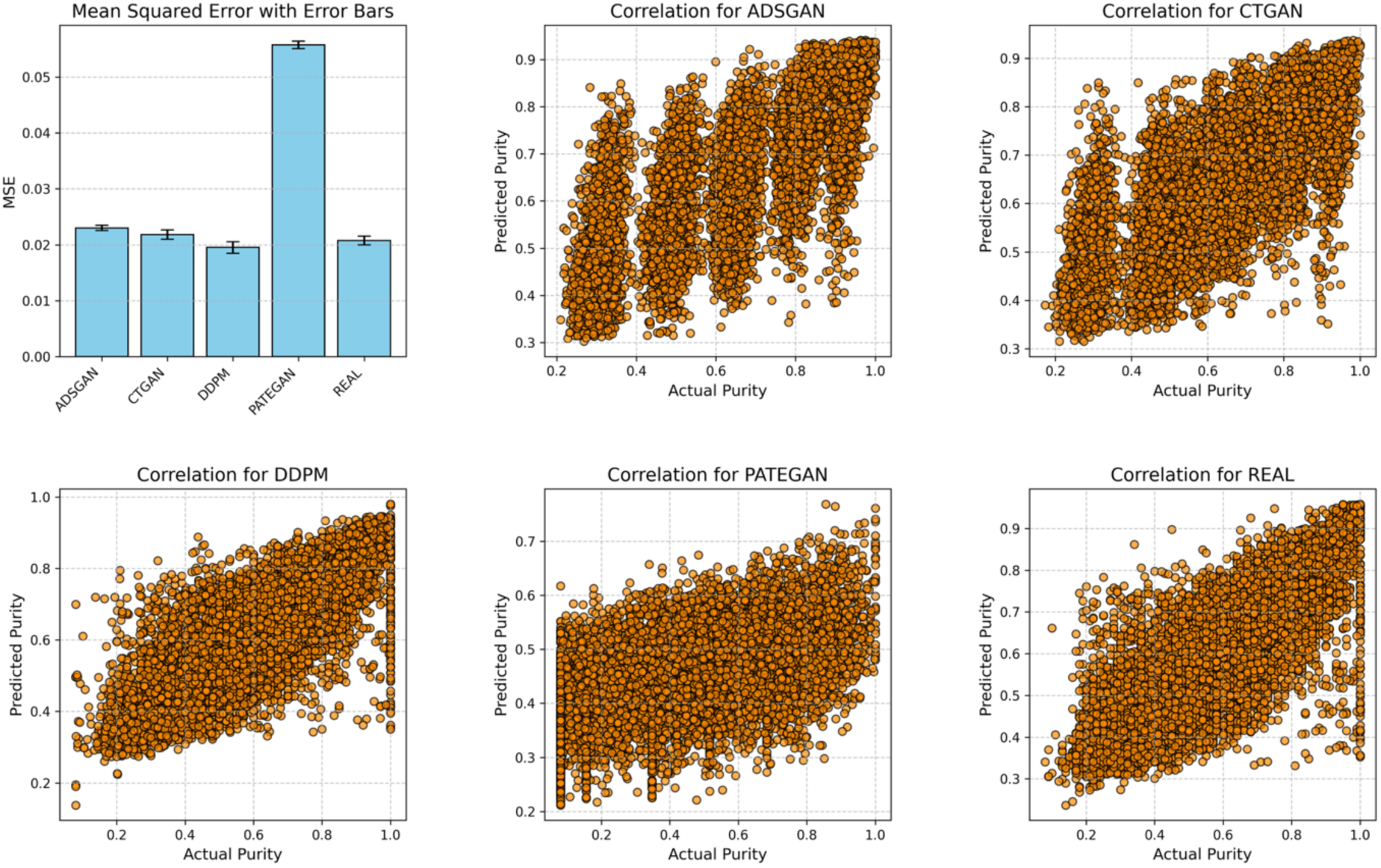
Scatter plots illustrate the correlation between actual and predicted tumour purity for the real dataset and four synthetic datasets. Abbreviations: ADSGAN, Anonymization through Data Synthesis using Generative Adversarial Networks; CT-GAN, conditional table generative adversarial network; DDPM, denoising diffusion probabilistic models; NHANES, National Health and Nutrition Examination Survey; PATEGAN, Private Aggregation of Teacher Ensembles Generative Adversarial Networks; MSE, mean squared error.

Augmentation – where synthetic data are added to the original data to reduce imbalances in particular features – is an important use-case for synthetic data. We thus asked SynthCraft to undertake ethnicity-specific augmentation of the original NHANES data. However, training predictive models of myocardial infarction on these augmented datasets did not improve either overall or sex- or ethnicity-specific discriminative performance (Figure 2).

Indeed, augmentation with synthetic data generated using PATEGAN, CTGAN, and DDPM led to reductions in the overall AUC and discriminative performance by subgroup by comparison with using the real data alone. Because the performance of models built on the original data were both high (AUC > 0.8) and consistent across subgroups, these findings are not out of the ordinary. But they highlight the need for iterative testing of synthetic data both across generators and use cases, and that synthetic data can balance representation across groups, but that does not necessarily remove bias or fairness^28^.

Our second use case for SynthCraft was in genomic data. Models trained on synthetic genomic data predict tumour purity. For the TCGA cohort, we generated five synthetic versions using PATE-GAN, ADS-GAN, CTGAN, and DDPM via the SynthCraft platform. Quality metrics for each synthetic dataset are summarized in Supplementary Table S4. We then compared the distributions of gene expression levels and tumour purity estimates between the real and synthetic cohorts, finding close alignment across all methods. To assess downstream analytical fidelity, we applied a previously identified gene signature^20^ in an XGBoost regression model to predict tumour purity. Predictive performance was comparable between the real data and all synthetic datasets except for PATE-GAN, which underperformed.

We repeated the same synthetic data generation pipeline for the TCGA cohort, demonstrating that SynthCraft can equally process high-dimensional genomic data. We found similar patterns to those in NHANES, where the statistical fidelity and utility of the synthetic data varied by generator (Supplementary Tables S4 and S5). XGBoost regression models predicting tumour purity had comparable mean squared errors when trained on synthetic data and the original TCGA cohort, except for models trained on synthetic data generated by PATEGAN (Figure 3).

## DISCUSSION

Despite its potential^2^, the use of synthetic data in practice is complex, from the selection and training of the synthetic data generator to context-specific evaluation. We demonstrate that an AI-based partner can support the systematic use of state-of-the-art synthetic data generation methods to develop and evaluate synthetic data across both tabular and genomic datasets entirely through natural language.

Improving the adoption, accessibility, reproducibility, and quality of research methods has relied upon improved training and a proliferation of research checklists and guidelines. These approaches are inherently limited: although guidelines can provide an approach to tackling a problem, there are few mechanisms to ensure their systematic use either by researchers or publishers^14^. Furthermore, it has been suggested that nearly 95% of time required to conduct analyses involving machine learning is spent programming, a technical debt^29^ that slows the dissemination of new methodological advances or potentially restricts access to teams with sufficient resources with contributing necessarily to scientific advance.

We show here that LLMs, specifically the use of agent-based frameworks, could support an alternative approach^30^ in which researchers interact with AI partners that encode and standardise good practice, ensuring that recommendations from guidelines and research checklists are considered, improving the quality of research performed. This approach simultaneously improves the accessibility of state-of-the-art ML methods: the user is no longer required to be an expert programmer, to be limited to synthetic data generators with which they have familiarity, or indeed to only those that are available in a single programming language.

Agent-based frameworks build on the potential for LLMs to act as reasoning engines^31^, with guardrails enforced through access only to pre-specified methodological software packages. This ensure that the resulting analyses are technically sound and is supported by the transparent reporting of any code run. A crucial feature of this approach is collaboration between the user and the framework – human-in-the-loop iteration – providing SynthCraft with domain knowledge to contextualise the problem; SynthCraft is a partner that augments a researcher, not a replacement.

Both SynthCraft and the underlying Synthcity package used to generate the synthetic data itself are open source, such that they verified, improved, or even adapted by the broader research community. As new synthetic data methods become available, these new tools can be incorporated into SynthCraft without requiring re-training of the underlying reasoning framework. We have demonstrated the creation of synthetic data for data access and augmentation, but SynthCraft is not limited to these, with any use case supported by the underlying Synthcity package. As with the underlying LLMs, Synthcity itself could be swapped for an alternative synthetic data generating package. SynthCraft has a specific purpose: the generation of synthetic data. This could be extended by linking other agent-based frameworks, for example to create downstream prediction models, which could operate in a cooperative manner to create end-to-end analytic pipelines.

SynthCraft has limitations. We found that the framework can requiring prompting to continue to the next stage of analyses, becoming focussed on the specific task at hand. With future research into training agent-based workflows and the ongoing improvement in underlying LLMs, we expect this to diminish. Although the underlying LLM does not have access inherently to the data, this safeguard could be bypassed by user prompting. Controlled instances of LLMs are available from cloud services that are compliant with healthcare privacy regulations and should be used to add security to the system; we provide instructions on how to set up SynthCraft with this feature built in.

In conclusion, we present an AI partner – SynthCraft – that enables the generation of synthetic datasets and augmented synthetic datasets using natural language. Such AI partners hold promise in democratising access to state-of-the-art methodologies, whilst improving the quality and reproducibility of analyses, furthering a new approach to scientific analysis.

## Data Availability

The data used for this study are publicly available online at

https://wwwn.cdc.gov/nchs/nhanes/continuousnhanes/default.aspx?Cycle=2021-2023

https://xenabrowser.net/datapages/?hub=https://tcga.xenahubs.net:443

## DATA SHARING STATEMENT

Both the NHANES and TCGA datasets are publicly available from the following links: NHANES - https://wwwn.cdc.gov/nchs/nhanes/continuousnhanes/default.aspx?Cycle=2021-2023 – and TCGA – https://xenabrowser.net/datapages/?hub=https://tcga.xenahubs.net:443.

The NHANES protocol was approved by the National Centre for Health Statistics (NCHS) Ethics Review Board. All NHANES participants provided written informed consent. All contributing sites to the TCGA had institutional review board approval to contribute specimens.

## CODE AVAILABILITY

SynthCraft is available from: https://github.com/vanderschaarlab/SynthCraft Synthcity is available from: https://github.com/vanderschaarlab/synthcity

## AUTHOR CONTRIBUTIONS

AB, MvdS, TC, RD and SRE conceived and designed the study. RD built SynthCraft. AB, TC, SRE and RD performed the analyses. TC, AB, SRE wrote the first draft of the manuscript. All authors contributed to reviewing and editing the manuscript. TC, AB, SRE, RD, MvdS had access to the datasets and take responsibility for the integrity of the analyses.

## ACKNOWLEDGEMENTS

The results shown here are in part based upon data generated by the TCGA Research Network: https://www.cancer.gov/tcga. We wish to thank all those who contributed as participants or researchers to the development of the NHANES and TCGA datasets. We are also indebted to William Weeks and the Microsoft AI for Good Research lab for their support and for Azure sponsorship credits.

