## Supplementary Appendix for "SynthCraft: an AI partner for synthetic data generation to support data access and augmentation in healthcare"

### SUPPLEMENTARY METHODS

#### Workflow of SynthCraft

SynthCraft integrates a reasoning engine that guides users through the process of synthetic data generation using a transparent, structured decision framework. Inspired by the CLiMB architecture[1], this reasoning process is formalised as an episodic multi-armed bandit[2], tailored to maximise utility (output quality) while minimising interaction cost (user burden).

The reasoning engine consists of several core components[1,2]:

- A set of tasks (**episodes**) necessary to complete an analysis plan
- **Costs** associated with specific actions that are returned as **feedback**
- **Actions** to complete the tasks, from invoking tools to asking the user for feedback
- A **state** that corresponds to all previous actions within the episode along with their costs

Let $E$ be the set of episode types or subtasks in the synthetic data pipeline, including:

1. Dataset intake and characterisation
2. Analytic intent elicitation
3. Synthetic data generation with or without specific privacy guarantees
4. Evaluation strategy selection
5. Iterative refinement
6. Transparent documentation

Note, data pre-processing is currently expected to be performed before data are passed to SynthCraft.

Each episode, $\rho$, corresponds to one subtask $e_\rho \in E$, and consists of a sequence of actions $(a_1^\rho, \ldots, a_t^\rho)$ drawn from a set of Actions, $\mathcal{A}$ [1,2]. These actions either progress the task or end the episode and engage the user. We enforce a maximum number of actions per episode before automatically discussing with the user to prevent infinite iteration[1].

The next action to be taken $a_t^\rho \in \mathcal{A}$ draws on the previous sequence of actions and the feedback received from each action[2]. Feedback is attributed a cost, $c$, of 0 or 1, and can be received from[1]:

- external tools (e.g. the results from statistical analyses; synthetic data generators);
- LLM self-reflection; or,
- the user themselves (either mid-episode or at the end of the episode).

Feedback that requires the user is penalised ($c = 1$) to minimise user burden. At the end of episode, there is a terminal reward $R^\rho$ again of 1 – corresponding to approval of the actions taken by SynthCraft – or 0 if the user requires the episode to be performed differently.

The reasoning agent maintains a dynamic plan over the subtasks. A subtask, $e_\rho$, is considered complete when the associated episode receives a reward $R^\rho = 1$. The plan is not strictly sequential; SynthCraft can reorder subtasks based on task context and user feedback, maintaining flexibility and robustness across different use cases. The ultimate objective of SynthCraft's reasoning system is to maximise terminal rewards by completing the analysis plan and its constituent steps efficiently with minimal user input[1]. All actions, feedback, and decisions are recorded to enable full reproducibility and auditability of the synthetic data workflow.

**Synthcity in-built four-stage augmentation pipeline**

Below are the four stages summarizing the synthetic data augmentation pipeline used to address class imbalances in ethnicity for the NHANES data.

***Stage 1: Baseline enumeration***
First the number of real participants in each self-reported ethnic subgroup are calculated. This audit revealed that several minority groups were markedly under-represented relative to the largest (majority) subgroup.

***Stage 2: Enrichment target calculation***
For each under-represented ethnicity, we computed an exact augmentation target - the number of additional records required to bring that subgroup's total up to parity with the majority group. This ensured that, once augmented, every ethnicity would appear equally often in downstream model training.

***Stage 3: Conditional synthetic generation***
We trained the synthetic generators on the full suite of NHANES variables. At synthesis time, the model was conditioned explicitly on the ethnicity label and the generator produced the requisite number of new records for each subgroup to better balance the ethnicity distribution in the dataset. Although entirely artificial, these samples preserved the multivariate relationships present in the original data.

***Stage 4: Cohort integration and evaluation***
Finally, the synthetic records were appended to the original NHANES data, yielding a balanced training set in which all ethnicities were equally represented. When classifiers and regression models were trained on this augmented cohort.

**Report generated on completing synthetic data generation for the NHANES dataset**

Let's summarize what has been done in the project so far:

1. **Upload Data File**:
    - The dataset nhanes_simple_v2.csv was successfully uploaded.
2. **Experiment Setup and Research Question**:
    - The user defined the research question and confirmed the classification task with the target column mcq160e.
3. **Exclude/Keep Columns**:
    - The column seqn was excluded from the analysis.
4. **Perform EDA**:
    - Exploratory Data Analysis (EDA) was performed on the dataset, revealing various statistical insights, correlations, and outliers.
5. **Generate Descriptive Statistics**:
    - Descriptive statistics were generated for the dataset.
6. **Confirm ML Problem Type**:
    - The ML problem type was confirmed to be classification.
7. **Select Synthetic Data Generation Method**:
    - The user chose the following synthetic data generators: CTGAN, TVAE, DDPM, ADSGAN, and PATEGAN.
8. **Generate Synthetic Data**:
    - Synthetic data was generated and augmented with the fairness column ridreth1.
9. **Perform EDA on Synthetic Data**:
    - EDA was performed on the synthetic dataset pategan_synthetic_data.csv.
10. **Generate Descriptive Statistics for Synthetic Data**:
    - Descriptive statistics were generated for the synthetic dataset.
11. **Plot Synthetic Data**:
    - Synthetic data was plotted for all selected methods, and the plots were saved in the working directory.

**Report generated on completing synthetic data generation for the TCGA dataset**

Let's summarize what has been done in the project so far:

1. **Upload Data File:**
    - The dataset named cleaned_gene_purity_data_v2.csv was uploaded.
2. **Experiment Setup and Research Question:**
    - The research problem was identified as a regression problem with the target column being "purity".
3. **Exclude/Keep Columns:**
    - The column "Unnamed: 0" was excluded from the dataset.

4. **Perform EDA:**
   - Exploratory Data Analysis (EDA) was performed on the dataset, revealing various statistics and correlations.
5. **Generate Descriptive Statistics:**
   - Descriptive statistics were generated for the dataset.
6. **Confirm ML Problem Type:**
   - The problem type was confirmed to be regression.
7. **Select Synthetic Data Generation Method:**
   - The user selected the following synthetic data generation methods: CTGAN, TVAE, DDPM, ADSGAN, and PATEGAN.
8. **Generate Synthetic Data:**
   - Synthetic data was generated successfully using the selected methods.
9. **Perform EDA on Synthetic Data:**
   - EDA was performed on the PATEGAN synthetic dataset.
10. **Generate Descriptive Statistics for Synthetic Data:**
    - Descriptive statistics were generated for the PATEGAN synthetic dataset.
11. **Plot Synthetic Data:**
    - Synthetic data plots were generated for all selected methods and saved in the working directory. diredirectory.

### SUPPLEMENTARY TABLES

**Supplementary Table S1. Summary of synthetic data generator tools available in SynthCraft**

| Generator | Description |
|---|---|
| **A) General purpose generators** | |
| Dummy sampler | Generates synthetic data by randomly sampling from real data. |
| Uniform sampler | Generates data by uniformly sampling across the variable range. |
| Marginal distributions | Samples each feature independently based on its marginal distribution. |
| CTGAN | GAN-based synthesizer tailored to tabular data with mixed types. |
| TVAE | Variational autoencoder adapted for tabular data. |
| Normalising flows | Neural normalizing flows for complex distribution modelling. |
| RTVAE | Recurrent VAE for longitudinal/time-series data. |
| Bayesian networks | Learns probabilistic graphical models from data. |
| GREAT | GAN with reinforcement learning-based generator optimization. |
| DDPM | Denoising diffusion probabilistic model for high-quality synthetic generation. |
| ARF | Adversarial random forest synthesizer using decision tree ensembles. |
| **B) Privacy-preserving generators** | |
| ADSGAN | Adversarially trains to fool re-identification attacks. |
| AIM | Minimizes mutual information between synthetic and real data. |
| PATEGAN | Differentially private GAN using the PATE framework. |
| DECAF | Privacy-preserving framework using attribute conditioning. |
| PrivBayes | Bayesian network generator with differential privacy guarantees. |
| DPGAN | GAN with gradient-level differential |

Summary of the general-purpose synthetic data generation methods (**A**) available in and privacy-preserving data generation methods (**B**) integrated into SynthCraft. The general-purpose generators include a range of statistical, adversarial, and deep generative models capable of modelling complex data distributions across both cross-sectional and temporal modalities. The privacy-preserving methods incorporate formal privacy mechanisms, such as differential privacy, mutual information minimization, and adversarial defence to reduce the risk of re-identification while maintaining utility in synthetic data.

**Supplementary Table S2. Data quality metrics in SynthCraft**

| Metric | Description |
|---|---|
| **A) Foundational aspects** | |
| Data mismatch | Measures structural incompatibility between real and synthetic datasets. |
| Common rows proportion | Proportion of synthetic rows identical to real rows (potential leakage). |
| Nearest synthetic neighbour distance | Distance from real samples to their closest synthetic neighbours. |
| Close values probability | Probability of synthetic samples closely matching real ones. |
| Distant values probability | Probability that synthetic data are unreasonably far from real samples. |
| **B) Statistical Similarity** | |
| Jensen-Shannon distance | Measures similarity between distributions via Jensen-Shannon divergence. |
| Chi-squared test | Compares categorical distributions using Chi-squared test. |
| Feature Corelation | Measures correlation preservation across features. |
| Kullback-Leibler divergence | KL divergence from synthetic to real (inverted). |
| Kolmogorov-Smirnov test | Kolmogorov-Smirnov test for univariate distribution similarity. |
| Empirical maximum mean discrepancy | Measures distributional distance using kernel-based two-sample test. |
| Wasserstein distance | Earth-mover's distance between real and synthetic samples. |
| Precision, Recall, Density & Coverage statistics | Precision/Recall metrics for distribution comparison using nearest neighbours. |
| Alpha precision (alpha precision, beta-recall, authenticity) | A 3-dimensional score of synthetic data quality consisting of measures of synthetic data fidelity (alpha-precision), diversity (beta-recall), and generalisability (authenticity) [3]. |
| Kaplan-Meier distance | Distance between real and synthetic Kaplan-Meier curves (for survival data). |
| **C) Downstream Performance**<br><br>Models are trained (linear regression, multi-layer perceptron [MLP], or XGBoost) on both real and synthetic data before they are evaluated on the real test data. $R^2$ is used for regression and the area under the receiver operating curve (AUC) for classification. | |
| Feature ranking distance | Compares feature importance rankings between real and synthetic-trained models. |
| **D) Privacy Risk Assessment** | |
| Delta presence | Measures the likelihood a real record exists in the synthetic set. |
| k-anonymization | Checks how many real samples are indistinguishable from others. |
| k-map | Assesses similarity of synthetic data to known real data populations. |
| Distinct l-diversity | Measures diversity of sensitive attributes within anonymized groups. |
| Identifiability score | Quantifies re-identification risk for individual records. |

Description of quality metrics implemented in SynthCraft; adapted from Synthcity[4], available at: https://github.com/vanderschaarlab/synthcity. Foundational aspects of synthetic data quality (**A**) related to those assessing data leakage, sample similarity, and basic distributional plausibility between real and synthetic datasets. The statistical similarity metrics **(B)** assess univariate and multivariate distributional alignment, dependency structure preservation, and overall fidelity of the synthetic data to the real data distribution. The downstream performance evaluation **(C)** metrics quantifies the utility of synthetic data. Models trained on synthetic data are compared to those trained on real data using various predictive algorithms, providing insight into the practical value of the generated datasets for real-world machine learning tasks. The privacy risk assessment metrics **(D)** assess individual-level disclosure risks, including re-identification, attribute inference, and sample uniqueness. Together, they provide a rigorous and interpretable evaluation of the privacy protections afforded by synthetic datasets.

**Supplementary Table S3. Performance metrics for generated synthetic datasets (NHANES)**

| Performance metric | Synthetic datasets | | | |
|---|---|---|---|---|
| | CTGAN | DDPM | ADSGAN | PATEGAN |
| Data mismatch ↓ | 0.0±0.0 | 0.0±0.0 | 0.0±0.0 | 0.0±0.0 |
| Common rows proportion ↓ | 0.0±0.0 | 0.0±0.0 | 0.0±0.0 | 0.0±0.0 |
| Nearest synthetic neighbour distance ↑ | 0.075±0.005 | 0.177±0.009 | 0.104±0.024 | 0.31±0.045 |
| Close values probability ↑ | 0.949±0.004 | 0.659±0.034 | 0.911±0.025 | 0.247±0.091 |
| Distant values probability ↓ | 0.013±0.002 | 0.002±0.001 | 0.023±0.002 | 0.01±0.004 |
| Jensen-Shannon distance ↓ | 0.012±0.001 | 0.007±0.001 | 0.011±0.001 | 0.016±0.002 |
| Chi-squared test ↑ | 0.477±0.043 | 0.809±0.071 | 0.474±0.04 | 0.915±0.04 |
| Kullback-Leibler divergence ↑ | 0.86±0.019 | 0.954±0.004 | 0.873±0.035 | 0.935±0.012 |
| Kolmogorov-Smirnov test ↑ | 0.886±0.013 | 0.946±0.008 | 0.904±0.006 | 0.86±0.02 |
| Empirical maximum mean discrepancy ↓ | 0.003±0.001 | 0.003±0.001 | 0.003±0.001 | 0.003±0.001 |
| Wasserstein distance ↓ | 0.058±0.003 | 0.159±0.006 | 0.054±0.003 | 0.295±0.066 |
| Precision ↑ | 0.977±0.008 | 0.953±0.007 | 0.986±0.002 | 0.811±0.056 |
| Recall ↑ | 0.957±0.016 | 0.983±0.002 | 0.942±0.013 | 0.992±0.003 |
| Density ↑ | 0.986±0.027 | 0.971±0.025 | 0.994±0.022 | 0.692±0.066 |
| Coverage ↑ | 0.843±0.01 | 0.964±0.006 | 0.848±0.029 | 0.899±0.007 |
| Alpha precision (one-class) ↑ | 0.903±0.036 | 0.914±0.02 | 0.894±0.019 | 0.791±0.037 |
| Beta-recall (one-class) ↑ | 0.404±0.011 | 0.464±0.015 | 0.407±0.018 | 0.224±0.02 |
| Authenticity (one-class) ↑ | 0.589±0.011 | 0.555±0.017 | 0.583±0.021 | 0.741±0.041 |
| (Performance) Linear model (train real test real) ↑ | 0.82±0.0 | 0.82±0.0 | 0.82±0.0 | 0.82±0.0 |
| (Performance) Linear model (train synthetic test synthetic) ↑ | 0.718±0.061 | 0.688±0.039 | 0.67±0.138 | 0.396±0.072 |
| (Performance) Linear model (train synthetic test real) ↑ | 0.635±0.088 | 0.655±0.115 | 0.653±0.081 | 0.255±0.029 |
| (Performance) MLP (train real test real) ↑ | 0.5±0.0 | 0.5±0.0 | 0.5±0.0 | 0.5±0.0 |
| (Performance) MLP (train synthetic test synthetic) ↑ | 0.5±0.0 | 0.5±0.0 | 0.5±0.0 | 0 |
| (Performance) MLP (train synthetic test real) ↑ | 0.5±0.0 | 0.5±0.0 | 0.5±0.0 | 0.5±0.0 |
| (Performance) XGBoost (train real test real) ↑ | 0.8±0.0 | 0.8±0.0 | 0.8±0.0 | 0.8±0.0 |
| (Performance) XGBoost (train synthetic test synthetic) ↑ | 0.632±0.016 | 0.704±0.059 | 0.641±0.078 | 0.5±0.072 |

**Supplementary Table S3. Performance metrics for generated synthetic datasets (NHANES)**

| Performance metric | Synthetic datasets | | | |
|---|---|---|---|---|
| | CTGAN | DDPM | ADSGAN | PATEGAN |
| (Performance) XGBoost (train synthetic test real) ↑ | 0.78±0.087 | 0.646±0.121 | 0.604±0.045 | 0.386±0.055 |
| (Performance) Linear model (train augmented test augmented) ↑ | 0.822±0.0 | 0.822±0.0 | 0.822±0.0 | 0.822±0.0 |
| (Performance) Linear model (train augmented test real) ↑ | 0.789±0.012 | 0.808±0.019 | 0.792±0.007 | 0.557±0.113 |
| (Performance) MLP (train augmented test augmented) ↑ | 0.5±0.0 | 0.5±0.0 | 0.5±0.0 | 0.5±0.0 |
| (Performance) MLP (train augmented test real) ↑ | 0.5±0.0 | 0.5±0.0 | 0.5±0.0 | 0.5±0.0 |
| (Performance) XGBoost (train augmented test augmented) ↑ | 0.743±0.001 | 0.743±0.001 | 0.743±0.001 | 0.743±0.001 |
| (Performance) XGBoost (train augmented test real) ↑ | 0.95±0.011 | 0.941±0.015 | 0.936±0.034 | 0.886±0.033 |
| Feature ranking distance (correlation) ↑ | 0.075±0.139 | 0.054±0.069 | 0.105±0.088 | 0.003±0.086 |
| Feature ranking distance (*P*-value) ↓ | 0.038±0.028 | 0.513±0.356 | 0.323±0.41 | 0.312±0.285 |
| Delta presence ↓ | 16.333±5.775 | 1.747±0.285 | 16.714±12.956 | 1.851±0.105 |
| k-anonymization (real data) ↑ | 28.0±0.0 | 28.0±0.0 | 28.0±0.0 | 28.0±0.0 |
| k-anonymization (synthetic data) ↑ | 12.333±0.471 | 20.667±8.34 | 13.667±3.3 | 23.667±4.922 |
| K-map ↑ | 3.333±0.943 | 23.667±0.943 | 5.667±5.249 | 22.0±3.266 |
| Distinct l-diversity (real data) ↑ | 28.0±0.0 | 28.0±0.0 | 28.0±0.0 | 28.0±0.0 |
| Distinct l-diversity (synthetic data) ↑ | 12.333±0.471 | 20.667±8.34 | 13.667±3.3 | 23.667±4.922 |
| Identifiability score ↓ | 0.426±0.008 | 0.495±0.021 | 0.428±0.02 | 0.256±0.02 |
| Identifiability score (one-class) ↓ | 0.398±0.02 | 0.462±0.017 | 0.401±0.015 | 0.235±0.036 |

An exhaustive list of statistical and performance metrics for the synthetic versions of NHANES by generator. Arrows refer to whether a higher or a lower score is desirable. Refer to Supplementary Table 2 or the SynthCity library in Github (https://github.com/vanderschaarlab/synthcity/) for further details. Abbreviations: ADSGAN, Anonymization through Data Synthesis using Generative Adversarial Networks; CT-GAN, conditional table generative adversarial network; DDPM, denoising diffusion probabilistic models; MLP, multilayer perceptron; NHANES, National Health and Nutrition Examination Survey; PATE-GAN, Private Aggregation of Teacher Ensembles Generative Adversarial Networks; XGBoost, Extreme Gradient Boosting.

**Supplementary Table S4. Exhaustive list of performance metrics for generated synthetic datasets (TCGA dataset)**

| Performance metric | Synthetic datasets | | | |
|---|---|---|---|---|
| | CTGAN | DDPM | ADSGAN | PATEGAN |
| Data mismatch ↓ | 0 ± 0 | 0 ± 0 | 0 ± 0 | 0 ± 0 |
| Common rows proportion ↓ | 0 ± 0 | 0 ± 0 | 0 ± 0 | 0 ± 0 |
| Nearest synthetic neighbour distance ↑ | 0.0328 ± 0.0185 | 0.0331 ± 0.0075 | 0.0131 ± 0.0021 | 0.0649 ± 0 |
| Close values probability ↑ | 0.9842 ± 0.0129 | 0.9835 ± 0.0066 | 0.9971 ± 0.0009 | 0.9607 ± 0 |
| Distant values probability ↓ | 0.001 ± 0.0004 | 0.0016 ± 0.0011 | 0.0009 ± 0.0002 | 0.0016 ± 0 |
| Jensen-Shannon distance ↓ | 0.0035 ± 0.0007 | 0.0015 ± 0.0001 | 0.0043 ± 0.0003 | 0.0073 ± 0 |
| Chi-squared test ↑ | 0.2222 ± 0.0393 | 0.1944 ± 0.0393 | 0.1389 ± 0.0393 | 0.8333 ± 0 |
| Kullback-Leibler divergence ↑ | 0.9518 ± 0.0134 | 0.9595 ± 0.0082 | 0.9274 ± 0.0041 | 0.954 ± 0 |
| Kolmogorov-Smirnov test ↑ | 0.8685 ± 0.0051 | 0.9643 ± 0.004 | 0.8599 ± 0.0205 | 0.8141 ± 0 |
| Empirical maximum mean discrepancy ↓ | 0.001 ± 0 | 0.001 ± 0 | 0.001 ± 0 | 0.001 ± 0 |
| Wasserstein distance ↓ | 0.0148 ± 0.0019 | 0.0188 ± 0.0046 | 0.0179 ± 0.0056 | 0.0995 ± 0 |
| Precision ↑ | 0.9415 ± 0.0192 | 0.9659 ± 0.0028 | 0.9499 ± 0.0084 | 0.7211 ± 0 |
| Recall ↑ | 0.9151 ± 0.0214 | 0.9559 ± 0.0061 | 0.9122 ± 0.0208 | 0.9845 ± 0 |
| Density ↑ | 0.9317 ± 0.0546 | 0.9959 ± 0.0081 | 0.9116 ± 0.0251 | 0.4616 ± 0 |
| Coverage ↑ | 0.834 ± 0.0549 | 0.9547 ± 0.0057 | 0.7996 ± 0.0457 | 0.5821 ± 0 |
| Alpha precision (one-class) ↑ | 0.7834 ± 0.0144 | 0.8493 ± 0.0053 | 0.8035 ± 0.0479 | 0.5306 ± 0 |
| Beta-recall (one-class) ↑ | 0.3979 ± 0.0198 | 0.4513 ± 0.0113 | 0.4179 ± 0.0094 | 0.2823 ± 0 |
| Authenticity (one-class) ↑ | 0.5229 ± 0.0085 | 0.5157 ± 0.0052 | 0.5367 ± 0.0082 | 0.6281 ± 0 |
| (Performance) Linear model (real data) $r^2$ ↑ | 0.2256 ± 0 | 0.2256 ± 0 | 0.2256 ± 0 | 0.2256 ± 0 |
| (Performance) Linear model (synthetic data) $r^2$ ↑ | 0.0896 ± 0.0941 | 0.1914 ± 0.0295 | 0.0166 ± 0.0826 | -0.7095 ± 0 |
| (Performance) Linear model (train synthetic test real) $r^2$ ↑ | 0.217 ± 0.0961 | 0.3162 ± 0.0062 | 0.2145 ± 0.0819 | -0.6887 ± 0 |
| (Performance) MLP (train real test real) $r^2$ ↑ | -10653.955 ± 0 | -10653.955 ± 0 | -10653.955 ± 0 | -10653.955 ± 0 |
| (Performance) MLP (train synthetic test synthetic) $r^2$ ↑ | -12318.803 ± 4326.696 | -18369.261 ± 12037.25 | -18778.1167 ± 7338.727 | -2365.9509 ± 0 |
| (Performance) MLP (train synthetic test real) $r^2$ ↑ | -23383.1862 ± 12249.21 | -17119.7328 ± 15873.54 | -24442.6468 ± 10756.83 | -2067.9063 ± 0 |
| (Performance) XGBoost (train real test real) $r^2$ ↑ | 0.3988 ± 0 | 0.3988 ± 0 | 0.3988 ± 0 | 0.3988 ± 0 |

**Supplementary Table S4. Exhaustive list of performance metrics for generated synthetic datasets (TCGA dataset)**

| Performance metric | Synthetic datasets | | | |
|---|---|---|---|---|
| | CTGAN | DDPM | ADSGAN | PATEGAN |
| (Performance) XGBoost (train synthetic test synthetic) $r^2$ ↑ | 0.2923 ± 0.0053 | 0.4151 ± 0.0026 | 0.2473 ± 0.1054 | -0.8171 ± 0 |
| (Performance) XGBoost (train synthetic test real) $r^2$ ↑ | 0.4099 ± 0.0351 | 0.4789 ± 0.0161 | 0.3304 ± 0.0897 | -0.804 ± 0 |
| Feature ranking distance (correlation) ↑ | 0.3455 ± 0.0297 | 0.1879 ± 0.1909 | 0.103 ± 0.1124 | 0.2364 ± 0 |
| Feature ranking distance (*P*-value) ↓ | 0.168 ± 0.0398 | 0.6071 ± 0.3988 | 0.6223 ± 0.186 | 0.3587 ± 0 |
| Delta presence ↓ | 4 ± 2.8284 | 3.3333 ± 0.9428 | 2.6618 ± 0.9658 | 1.4114 ± 0 |
| k-anonymization (real data) ↑ | 2 ± 0 | 2 ± 0 | 2 ± 0 | 2 ± 0 |
| k-anonymization (synthetic data) ↑ | 1.6667 ± 0.9428 | 1 ± 0 | 4 ± 2.4495 | 6 ± 0 |
| K-map ↑ | 1.3333 ± 0.4714 | 1.3333 ± 0.4714 | 5.3333 ± 3.6818 | 5 ± 0 |
| Distinct l-diversity (real data) ↑ | 2 ± 0 | 2 ± 0 | 2 ± 0 | 2 ± 0 |
| Distinct l-diversity (synthetic data) ↑ | 1.6667 ± 0.9428 | 1 ± 0 | 4 ± 2.4495 | 6 ± 0 |
| Identifiability score ↓ | 0.3764 ± 0.0256 | 0.4761 ± 0.0124 | 0.3583 ± 0.0135 | 0.1911 ± 0 |
| Identifiability score (one-class) ↓ | 0.4268 ± 0.014 | 0.4747 ± 0.0081 | 0.4291 ± 0.022 | 0.2438 ± 0 |

Performance metrics for the different synthetic versions generated of the TCGA dataset. Arrows indicate whether a higher or lower value is preferable. Abbreviations: ADSGAN, Anonymization through Data Synthesis using Generative Adversarial Networks; CT-GAN, conditional table generative adversarial network; DDPM, denoising diffusion probabilistic models; MLP, multilayer perceptron; NHANES, National Health and Nutrition Examination Survey; PATE-GAN, Private Aggregation of Teacher Ensembles Generative Adversarial Networks; XGBoost, Extreme Gradient Boosting.

**Supplementary Table S5. Comparison of variable distributions in the real and synthetically generated datasets (TCGA purity dataset)**

| Variable | Real dataset | Synthetic datasets | | | |
|---|---|---|---|---|---|
| | | ADSGAN | CT-GAN | DDPM | PATE-GAN |
| **C1S** | 8150 ± 14220 | 7345 ± 12219 | 7355 ± 12448 | 7843 ± 13246 | 11703 ± 21505 |
| **CCDC69** | 630 ± 1049 | 483 ± 782 | 600 ± 945 | 598 ± 1224 | 1823 ± 3364 |
| **CCL21** | 578 ± 3560 | 573 ± 2720 | 800 ± 5316 | 787 ± 7900 | 2266 ± 8856 |
| **CCL22** | 127 ± 396 | 161 ± 390 | 146 ± 474 | 129 ± 521 | 465 ± 1311 |
| **CSF2RB** | 312 ± 575 | 257 ± 422 | 333 ± 644 | 312 ± 695 | 574 ± 1371 |
| **CYTIP** | 239 ± 345 | 248 ± 340 | 261 ± 362 | 237 ± 394 | 331 ± 650 |
| **FGR** | 227 ± 279 | 190 ± 232 | 207 ± 226 | 224 ± 304 | 412 ± 684 |
| **IL7R** | 118 ± 209 | 114 ± 198 | 77 ± 151 | 118 ± 235 | 166 ± 303 |
| **POU2AF1** | 296 ± 1043 | 343 ± 1087 | 477 ± 1758 | 361 ± 2032 | 724 ± 2638 |
| **RHOH** | 180 ± 420 | 233 ± 485 | 244 ± 520 | 166 ± 407 | 220 ± 494 |
| **Purity** | 0.625 ± 0.209 | 0.658 ± 0.211 | 0.655 ± 0.201 | 0.631 ± 0.206 | 0.456 ± 0.248 |

The set of genes presented was selected from Li et al. (2019)[5] and tumour purity was derived using the ABSOLUTE method[6].

All data are mean (± standard deviation). The distribution of variables in the original selection of data from the TCGA purity dataset (i.e., the "real" dataset) and four synthetic datasets from various methods are given. Abbreviations: ADSGAN, Anonymization through Data Synthesis using Generative Adversarial Networks; CT-GAN, conditional table generative adversarial network; DDPM, denoising diffusion probabilistic models; PATE-GAN, Private Aggregation of Teacher Ensembles Generative Adversarial Networks

.

**Supplementary Table S6. Performance on the TCGA gene purity dataset**

| Method | MSE (mean ± SD) |
|---|---|
| Real data | 0.0208 ± 0.0008 |
| ADSGAN | 0.0230 ± 0.0005 |
| CT-GAN | 0.0218 ± 0.0008 |
| DDPM | 0.0195 ± 0.0010 |
| PATE-GAN | 0.0558 ± 0.0007 |
| TVAE | 0.0242 ± 0.0010 |

XGBoost performance predicting the tumour purity in bulk RNA samples using a previously derived prognostic gene expression signature[5,6]. Results are averaged across 5-folds for each method and reported with the corresponding standard deviation. Abbreviations: ADS-GAN, Anonymization through Data Synthesis using Generative Adversarial Networks; CT-GAN, conditional table generative adversarial network; DDPM, denoising diffusion probabilistic models; PATE-GAN, Private Aggregation of Teacher Ensembles Generative Adversarial Networks

.

### SUPPLEMENTARY FIGURES

#### Supplementary Figure S1. Workflow for the NHANES dataset

The following set of screenshots, illustrate the user-AI interaction when using SynthCraft.

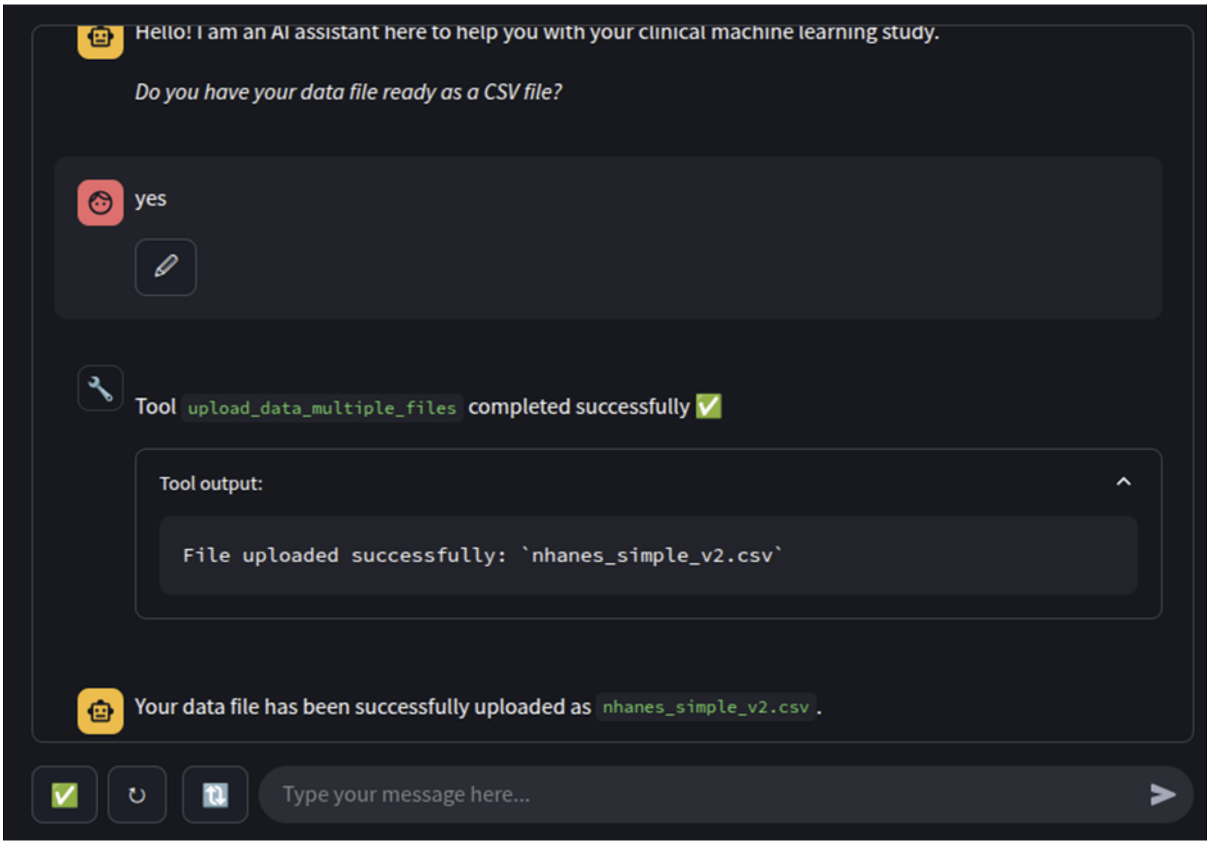

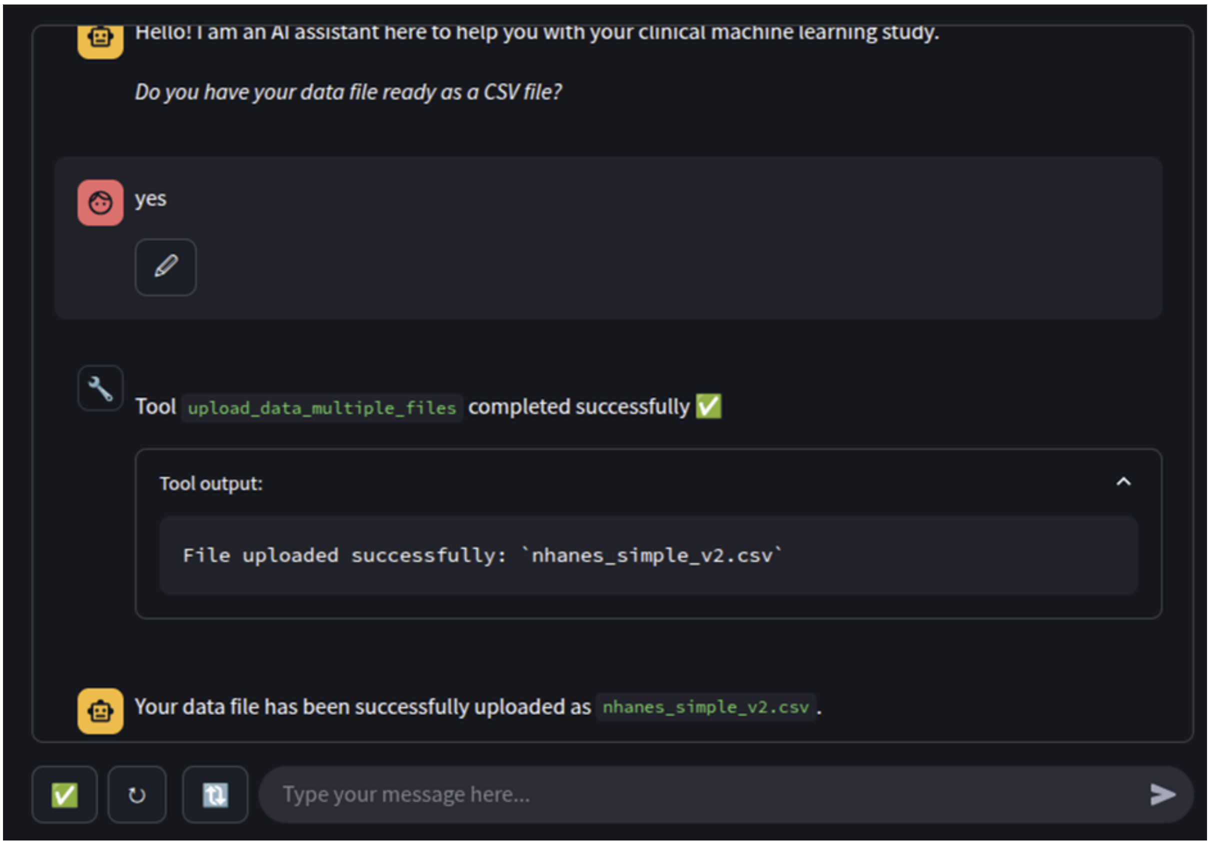
Hello! I am an AI assistant here to help you with your clinical machine learning study.
Do you have your data file ready as a CSV file?
yes
Tool upload_data_multiple_files completed successfully ✅
Tool output:
File uploaded successfully: `nhanes_simple_v2.csv`
Your data file has been successfully uploaded as nhanes_simple_v2.csv.
Type your message here...

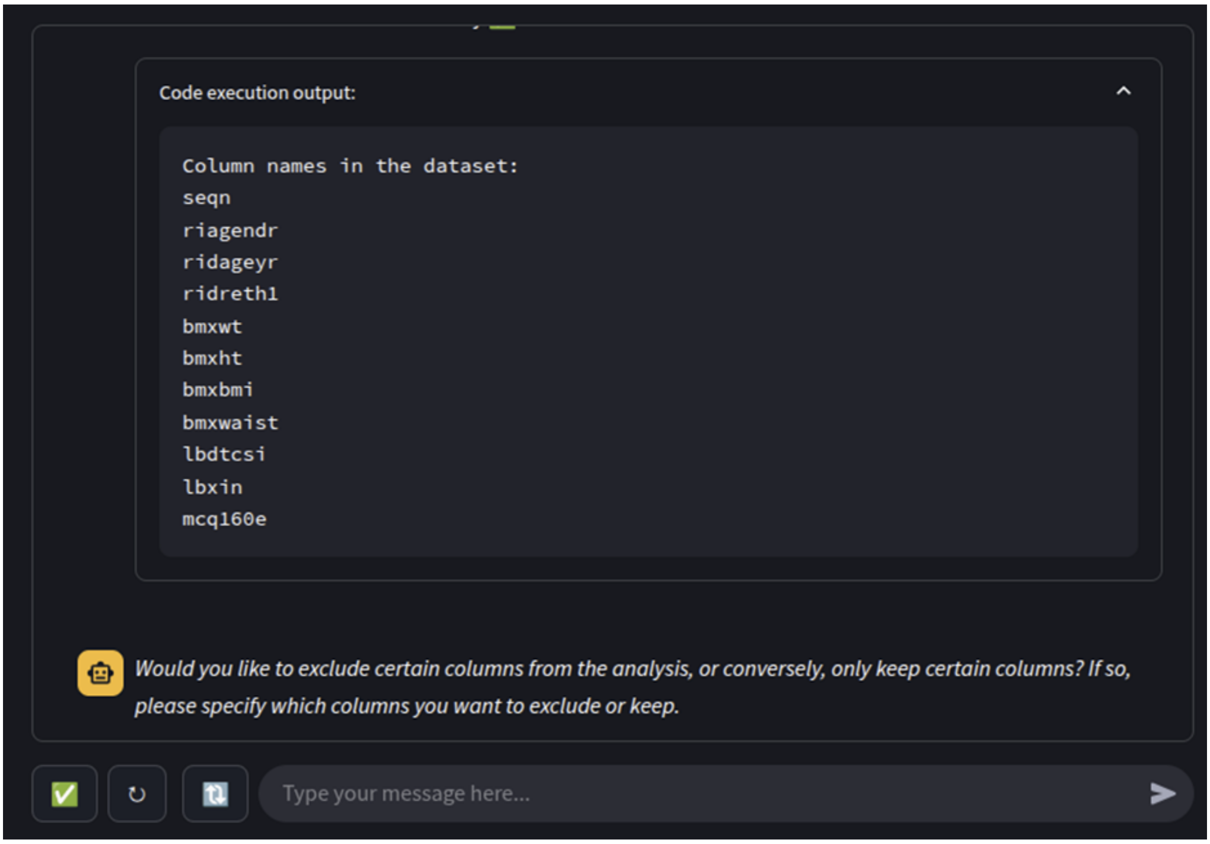
Code execution output:
Column names in the dataset:
seqn
riagendr
ridageyr
ridreth1
bmxwt
bmxht
bmxbmi
bmxwaist
lbdtcsi
lbxin
mcq160e
Would you like to exclude certain columns from the analysis, or conversely, only keep certain columns? If so, please specify which columns you want to exclude or keep.
Type your message here...

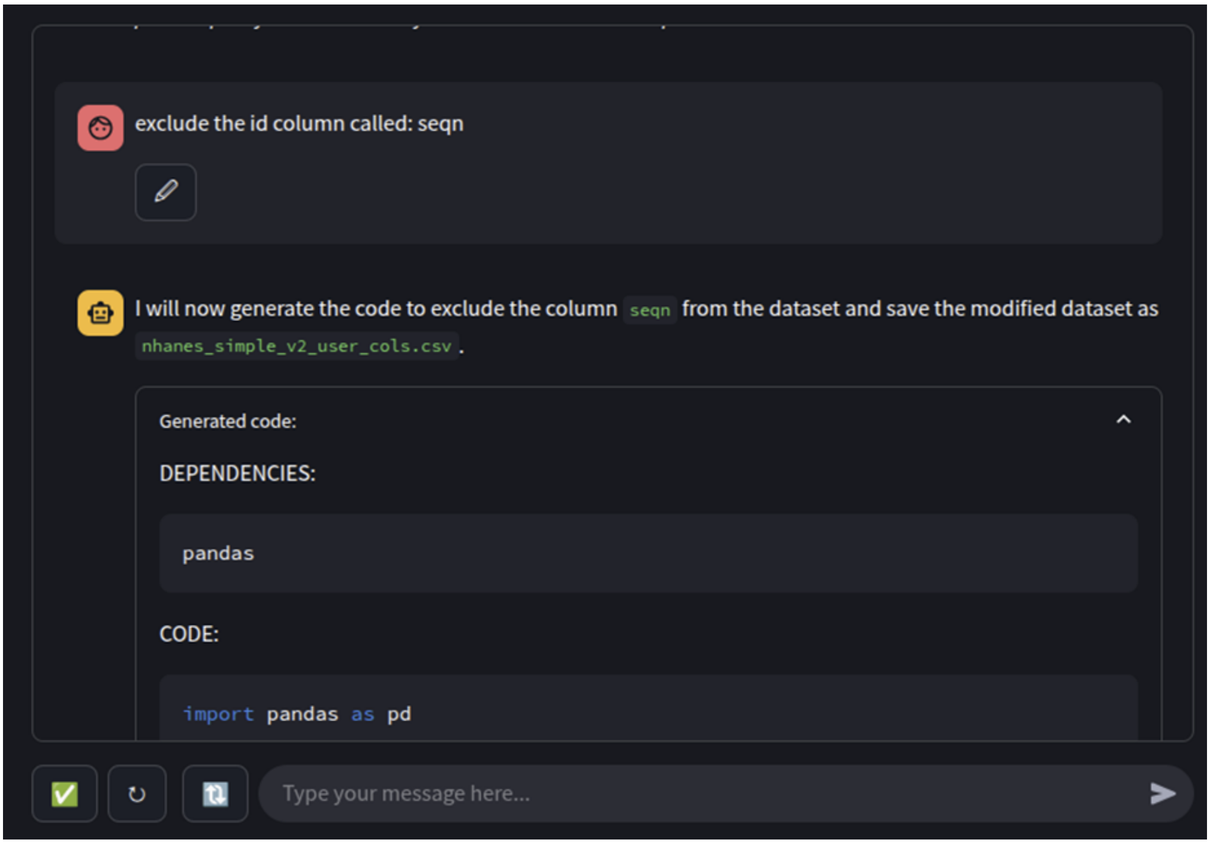
exclude the id column called: seqn
I will now generate the code to exclude the column seqn from the dataset and save the modified dataset as nhanes_simple_v2_user_cols.csv .
Generated code:
DEPENDENCIES:
pandas
CODE:
import pandas as pd
Type your message here...

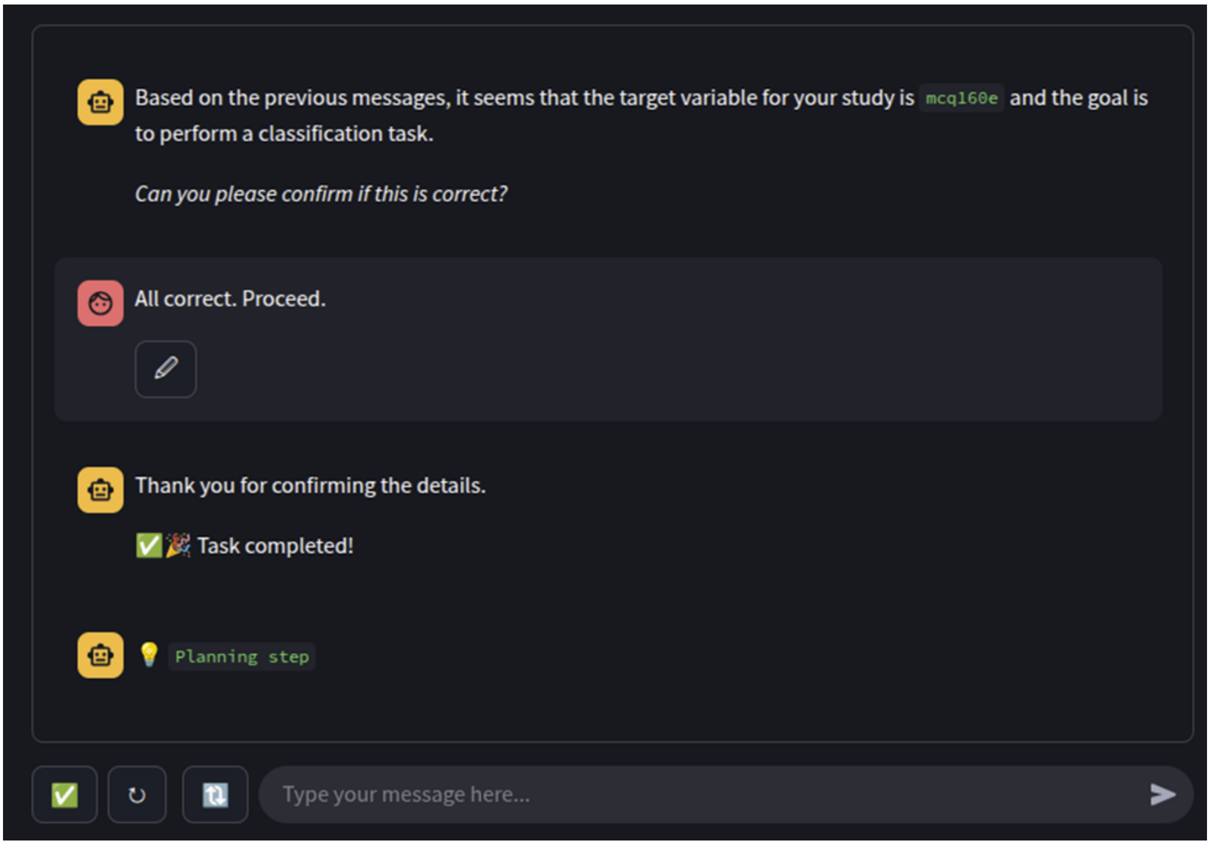
Based on the previous messages, it seems that the target variable for your study is mcq160e and the goal is to perform a classification task.
Can you please confirm if this is correct?
All correct. Proceed.
Thank you for confirming the details.
✅🎉 Task completed!
💡 Planning step
Type your message here...

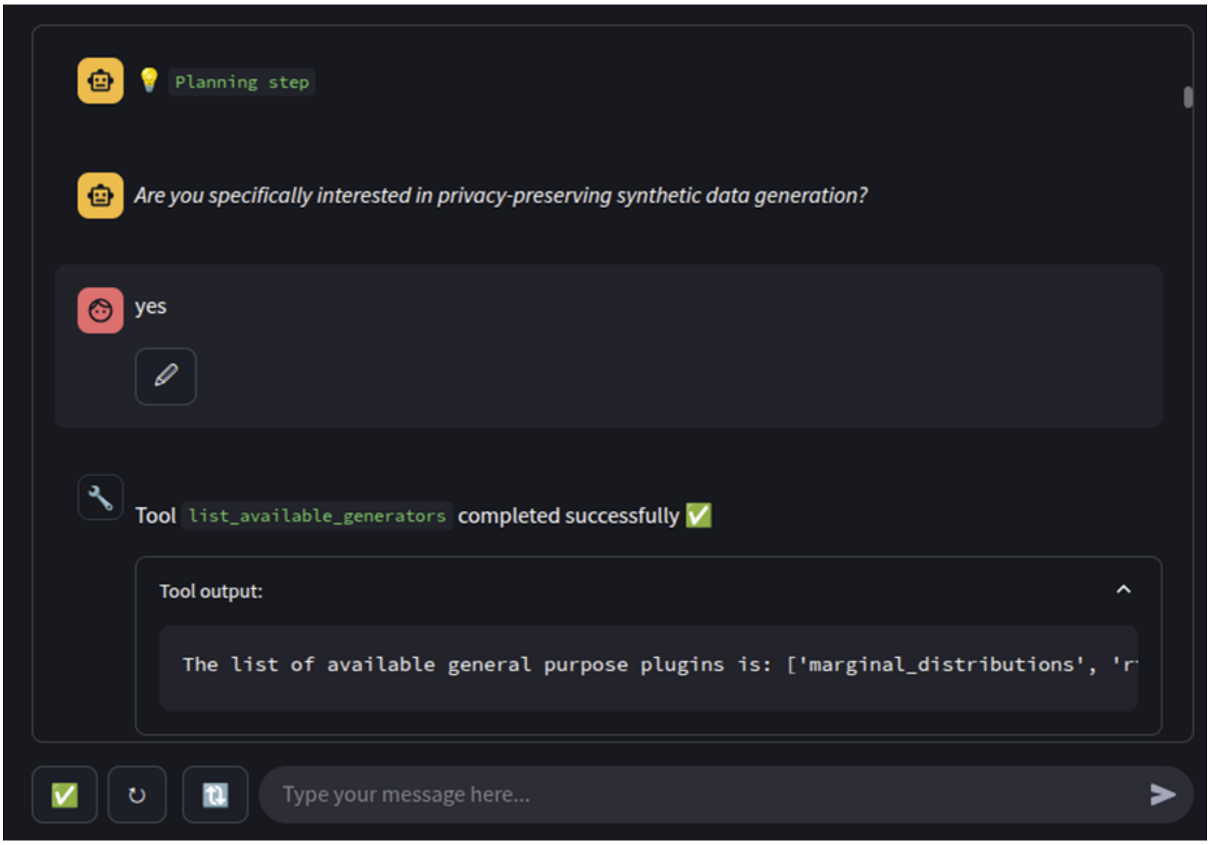
Planning step
Are you specifically interested in privacy-preserving synthetic data generation?
yes
Tool list_available_generators completed successfully
Tool output:
The list of available general purpose plugins is: ['marginal_distributions', 'r
Type your message here...

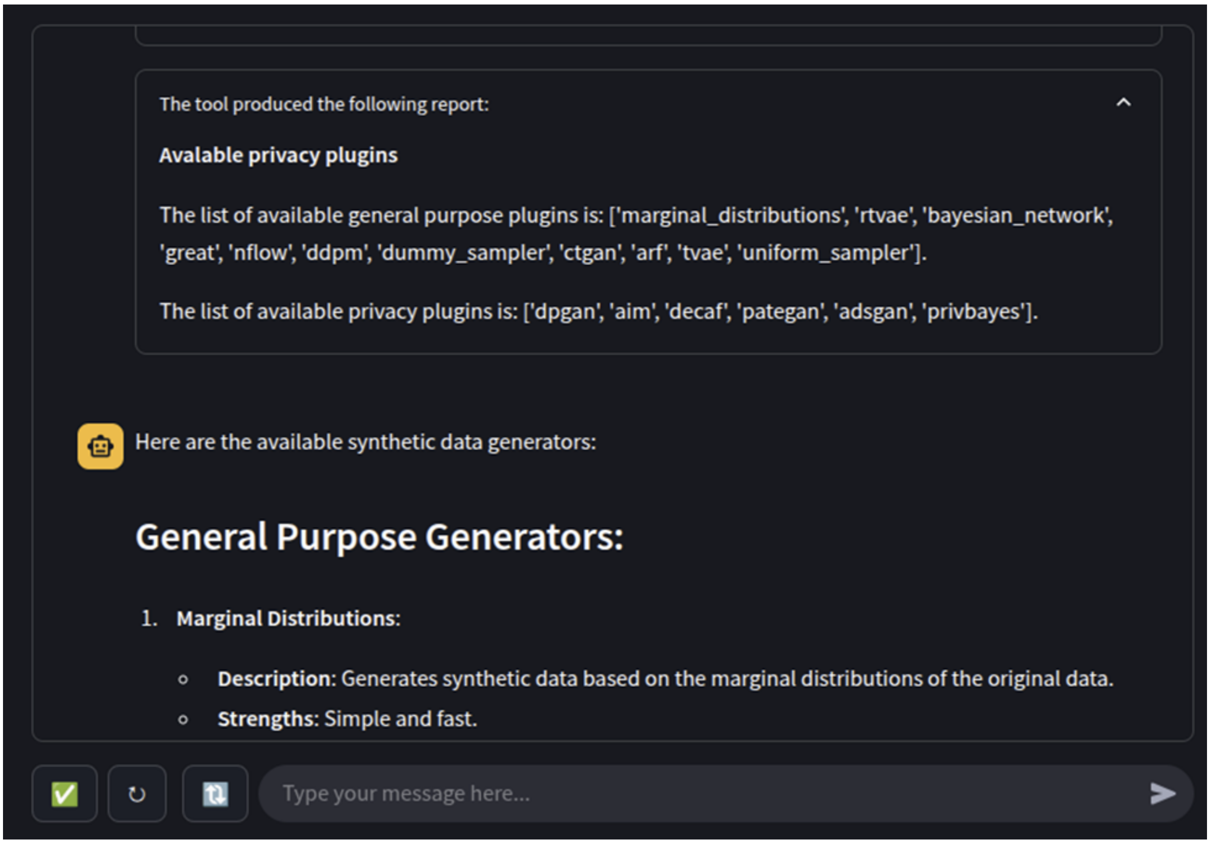
The tool produced the following report:
Avalable privacy plugins
The list of available general purpose plugins is: ['marginal_distributions', 'rtvae', 'bayesian_network', 'great', 'nflow', 'ddpm', 'dummy_sampler', 'ctgan', 'arf', 'tvae', 'uniform_sampler'].
The list of available privacy plugins is: ['dpgan', 'aim', 'decaf', 'pategan', 'adsgan', 'privbayes'].
Here are the available synthetic data generators:
General Purpose Generators:
1. Marginal Distributions:
Description: Generates synthetic data based on the marginal distributions of the original data.
Strengths: Simple and fast.
Type your message here...

- **Strengths:** Simple and fast.
- **Weaknesses:** Does not capture dependencies between variables.

2. **RTVAE:**
   - **Description:** Recurrent Tabular Variational Autoencoder, generates synthetic data using a deep learning model.
   - **Strengths:** Captures complex dependencies.
   - **Weaknesses:** Requires significant computational resources.
3. **Bayesian Network:**
   - **Description:** Uses Bayesian networks to generate synthetic data.
   - **Strengths:** Captures probabilistic relationships between variables.
   - **Weaknesses:** Can be computationally intensive and requires expert knowledge.
4. **GREAT:**
   - **Description:** Generates synthetic data using a graph-based approach.
   - **Strengths:** Captures complex relationships.
   - **Weaknesses:** Can be computationally intensive.
5. **NFlow:**

✅ ↻ 🔃 Type your message here...

[illegible]

- **Weaknesses:** Requires significant computational resources.

5. **ADSGAN:**
   - **Description:** GDPR-like privacy engine for synthetic data generation.
   - **Strengths:** Provides GDPR-like privacy guarantees.
   - **Weaknesses:** Can be computationally intensive.
6. **PrivBayes:**
   - **Description:** Privacy-preserving synthetic data generation using Bayesian networks.
   - **Strengths:** Provides privacy guarantees.
   - **Weaknesses:** Can be computationally intensive.

*Please provide a list of the generators you would like to use from the above options.*

adsgan, ctgan, arf, ddpm

✅ ↻ 🔃 Type your message here...

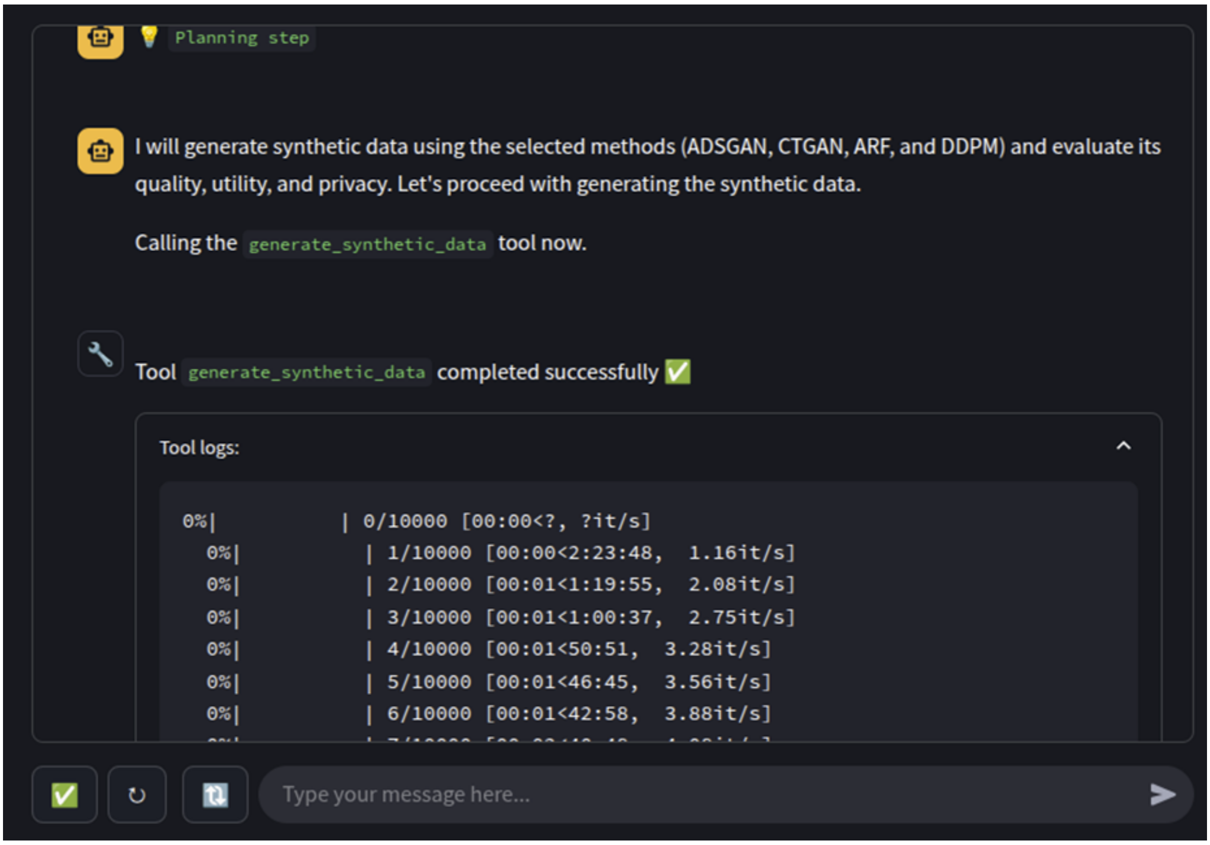
Planning step
I will generate synthetic data using the selected methods (ADSGAN, CTGAN, ARF, and DDPM) and evaluate its quality, utility, and privacy. Let's proceed with generating the synthetic data.
Calling the generate_synthetic_data tool now.
Tool generate_synthetic_data completed successfully ✅
Tool logs:
0%| | 0/10000 [00:00<?, ?it/s]
0%| | 1/10000 [00:00<2:23:48, 1.16it/s]
0%| | 2/10000 [00:01<1:19:55, 2.08it/s]
0%| | 3/10000 [00:01<1:00:37, 2.75it/s]
0%| | 4/10000 [00:01<50:51, 3.28it/s]
0%| | 5/10000 [00:01<46:45, 3.56it/s]
0%| | 6/10000 [00:01<42:58, 3.88it/s]
Type your message here...

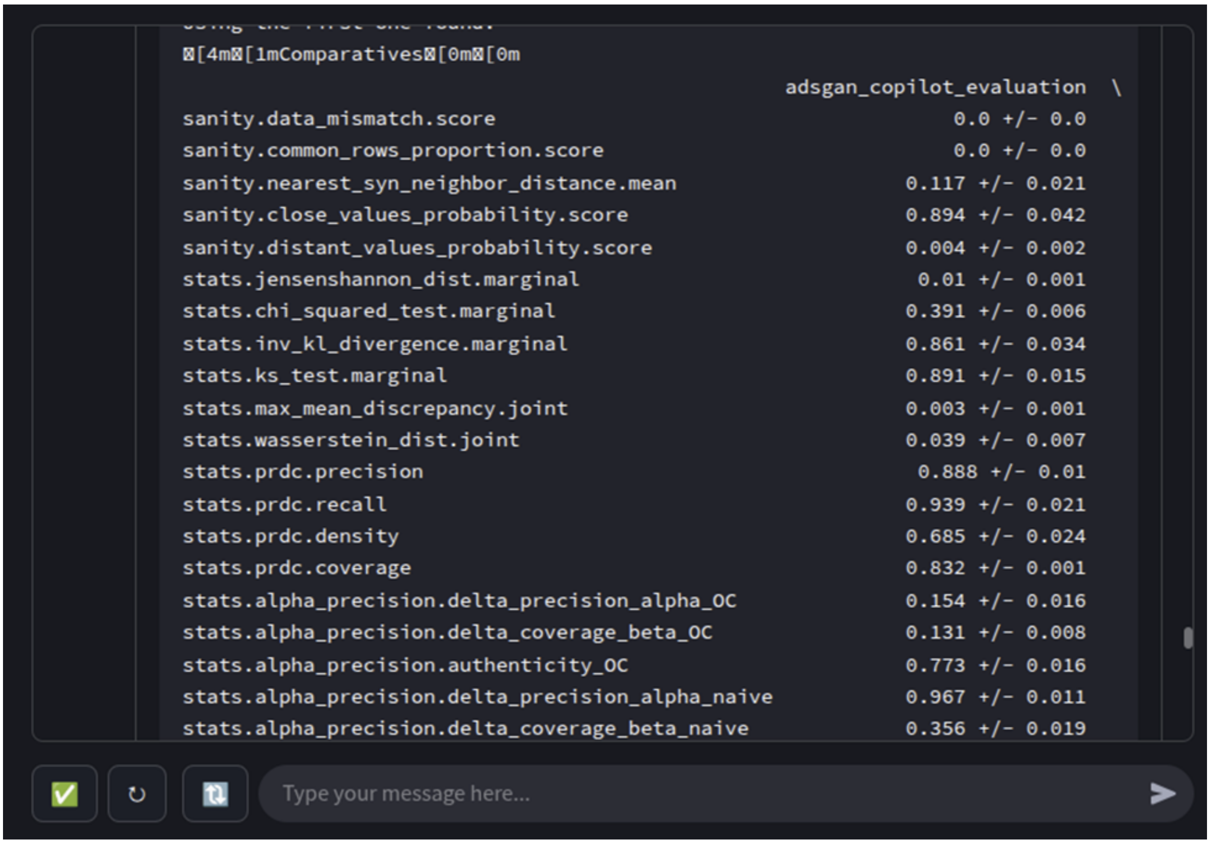
⊠[4m⊠[1mComparatives⊠[0m⊠[0m
adsgan_copilot_evaluation \
sanity.data_mismatch.score 0.0 +/- 0.0
sanity.common_rows_proportion.score 0.0 +/- 0.0
sanity.nearest_syn_neighbor_distance.mean 0.117 +/- 0.021
sanity.close_values_probability.score 0.894 +/- 0.042
sanity.distant_values_probability.score 0.004 +/- 0.002
stats.jensenshannon_dist.marginal 0.01 +/- 0.001
stats.chi_squared_test.marginal 0.391 +/- 0.006
stats.inv_kl_divergence.marginal 0.861 +/- 0.034
stats.ks_test.marginal 0.891 +/- 0.015
stats.max_mean_discrepancy.joint 0.003 +/- 0.001
stats.wasserstein_dist.joint 0.039 +/- 0.007
stats.prdc.precision 0.888 +/- 0.01
stats.prdc.recall 0.939 +/- 0.021
stats.prdc.density 0.685 +/- 0.024
stats.prdc.coverage 0.832 +/- 0.001
stats.alpha_precision.delta_precision_alpha_OC 0.154 +/- 0.016
stats.alpha_precision.delta_coverage_beta_OC 0.131 +/- 0.008
stats.alpha_precision.authenticity_OC 0.773 +/- 0.016
stats.alpha_precision.delta_precision_alpha_naive 0.967 +/- 0.011
stats.alpha_precision.delta_coverage_beta_naive 0.356 +/- 0.019
Type your message here...

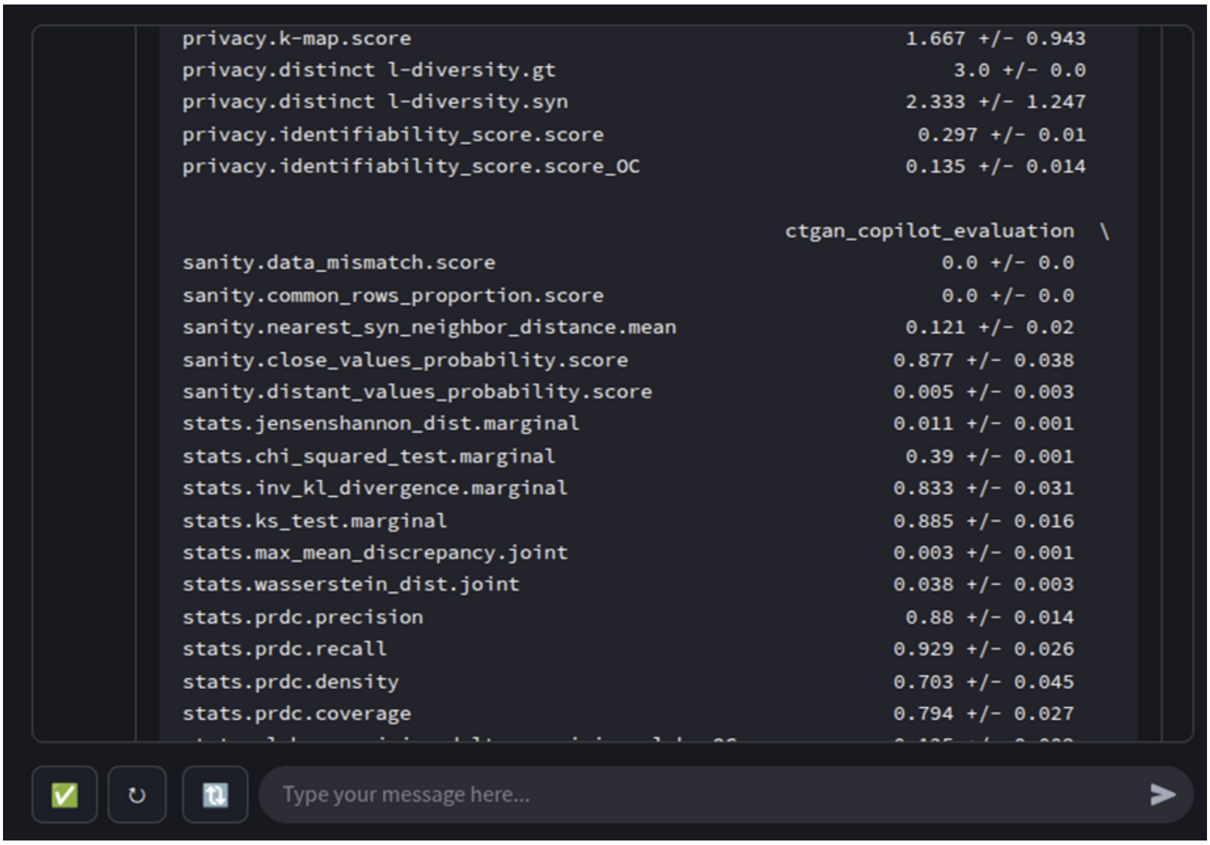
privacy.k-map.score 1.667 +/- 0.943
privacy.distinct l-diversity.gt 3.0 +/- 0.0
privacy.distinct l-diversity.syn 2.333 +/- 1.247
privacy.identifiability_score.score 0.297 +/- 0.01
privacy.identifiability_score.score_OC 0.135 +/- 0.014
ctgan_copilot_evaluation \
sanity.data_mismatch.score 0.0 +/- 0.0
sanity.common_rows_proportion.score 0.0 +/- 0.0
sanity.nearest_syn_neighbor_distance.mean 0.121 +/- 0.02
sanity.close_values_probability.score 0.877 +/- 0.038
sanity.distant_values_probability.score 0.005 +/- 0.003
stats.jensenshannon_dist.marginal 0.011 +/- 0.001
stats.chi_squared_test.marginal 0.39 +/- 0.001
stats.inv_kl_divergence.marginal 0.833 +/- 0.031
stats.ks_test.marginal 0.885 +/- 0.016
stats.max_mean_discrepancy.joint 0.003 +/- 0.001
stats.wasserstein_dist.joint 0.038 +/- 0.003
stats.prdc.precision 0.88 +/- 0.014
stats.prdc.recall 0.929 +/- 0.026
stats.prdc.density 0.703 +/- 0.045
stats.prdc.coverage 0.794 +/- 0.027
Type your message here...

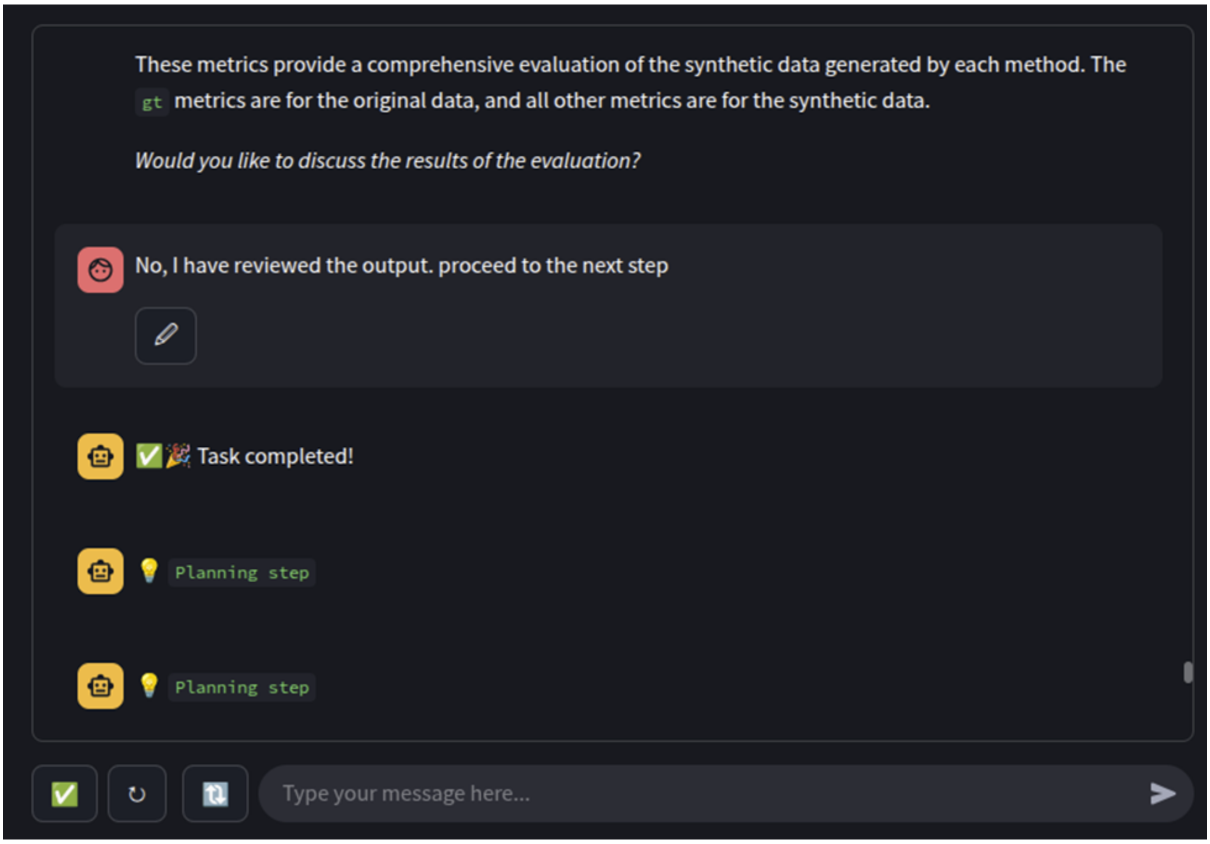
These metrics provide a comprehensive evaluation of the synthetic data generated by each method. The gt metrics are for the original data, and all other metrics are for the synthetic data.
Would you like to discuss the results of the evaluation?
No, I have reviewed the output. proceed to the next step
✅🎉 Task completed!
💡 Planning step
💡 Planning step
Type your message here...

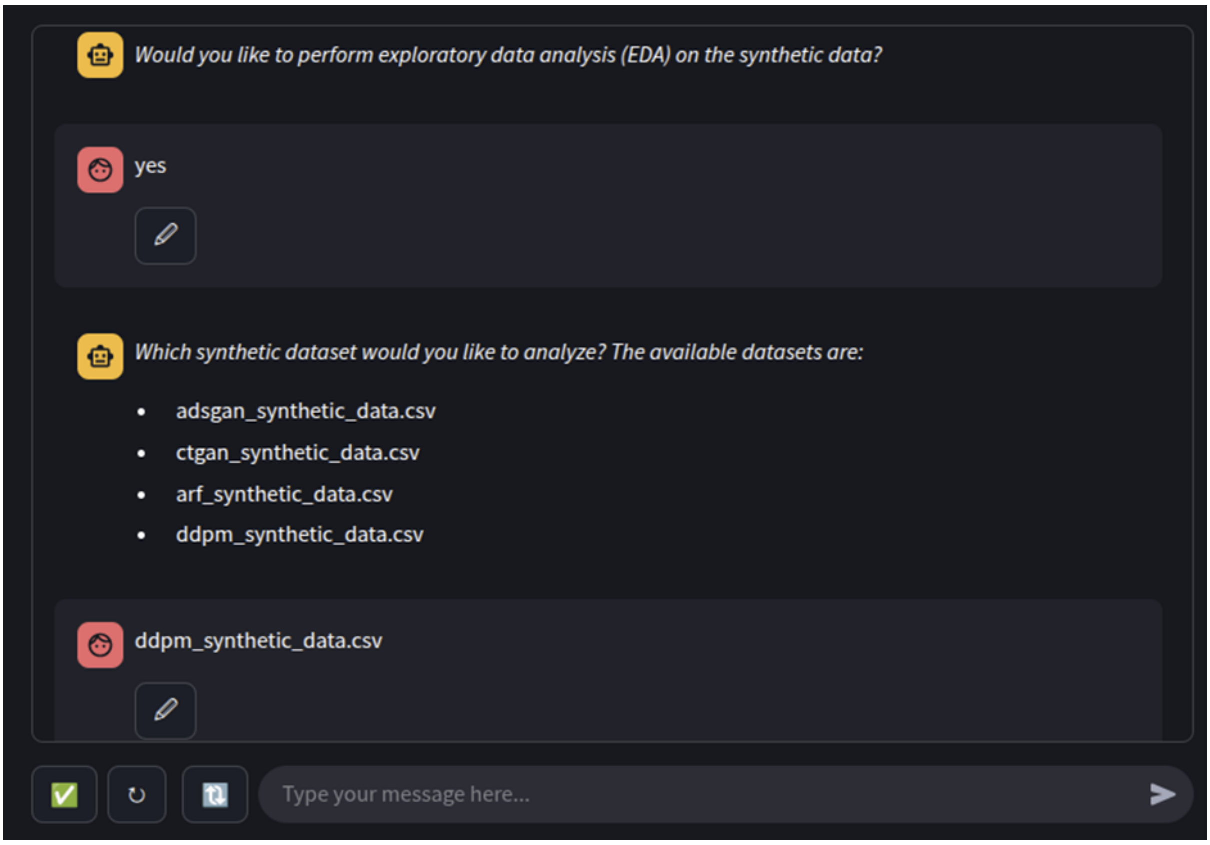
Would you like to perform exploratory data analysis (EDA) on the synthetic data?
yes
Which synthetic dataset would you like to analyze? The available datasets are:
adsgan_synthetic_data.csv
ctgan_synthetic_data.csv
arf_synthetic_data.csv
ddpm_synthetic_data.csv
ddpm_synthetic_data.csv
Type your message here...
